# An international survey on the impact of COVID-19 in individuals with Down syndrome

**DOI:** 10.1101/2020.11.03.20225359

**Authors:** Anke Hüls, Alberto C. S. Costa, Mara Dierssen, R. Asaad Baksh, Stefania Bargagna, Nicole T. Baumer, Ana Claudia Brandão, Angelo Carfi, Maria Carmona-Iragui, Brain Allen Chicoine, Sujay Ghosh, Monica Lakhanpaul, Coral Manso, Miguel-Angel Mayer, Maria del Carmen Ortega, Diego Real de Asua, Anne-Sophie Rebillat, Lauren Ashley Russell, Giuseppina Sgandurra, Diletta Valentini, Stephanie L Sherman, Andre Strydom, on behalf of the T21RS COVID-19 Initiative

## Abstract

**Background:** Health conditions and immune dysfunction associated with trisomy 21 (Down syndrome, DS) may impact the clinical course of COVID-19 once infected by SARS-CoV-2.

**Methods:** The T21RS COVID-19 Initiative launched an international survey for clinicians or caregivers/family members on patients with COVID-19 and DS (N=1046). De-identified survey data collected between April and October 2020 were analysed and compared with the UK ISARIC4C survey of hospitalized COVID-19 patients with and without DS. COVID-19 patients with DS from the ISARIC4C survey (ISARIC4C DS cases=100) were matched to a random set of patients without DS (ISARIC4C controls=400) and hospitalized DS cases in the T21RS survey (T21RS DS cases=100) based on age, gender, and ethnicity.

**Findings:** The mean age in the T21RS survey was 29 years (SD=18), 73% lived with their family. Similar to the general population, the most frequent signs and symptoms of COVID-19 were fever, cough, and shortness of breath. Pain and nausea were reported less frequently (p<0.01), whereas altered consciousness/confusion were reported more frequently (p<0.01). Risk factors for hospitalization and mortality were similar to the general population (age, male gender, diabetes, obesity, dementia) with the addition of congenital heart defects as a risk factor for hospitalization. Mortality rates showed a rapid increase from age 40 and were higher than for controls (T21RS DS versus controls: risk ratio (RR)=3.5 (95%-CI=2.6;4.4), ISARIC4C DS versus controls: RR=2.9 (95%-CI=2.1;3.8)) even after adjusting for known risk factors for COVID-19 mortality.

**Interpretation:** Leading signs/symptoms of COVID-19 and risk factors for severe disease course are similar to the general population. However, individuals with DS present significantly higher rates of mortality, especially from age 40.

**Funding:** Down Syndrome Affiliates in Action, Down Syndrome Medical Interest Group-USA, GiGi’s Playhouse, Jerome Lejeune Foundation, LuMind IDSC Foundation, Matthews Foundation, National Down Syndrome Society, National Task Group on Intellectual Disabilities and Dementia Practices.

## INTRODUCTION

Down syndrome (DS), the result of the trisomy of chromosome 21, is the commonest genetic cause for intellectual disability. It is associated with specific co-occurring health conditions and immune dysfunction that may impact the clinical course and risk for life-threatening disease due to infection by the novel severe acute respiratory syndrome coronavirus 2 (SARS-CoV-2) ^1^.

Immune-response dysfunction associated with trisomy 21 includes alterations in the number of different types of immune cells (T- and B-cells, monocytes and neutrophils) ^2^ and adverse antibody responses ^3^. The unusual inflammatory profile of individuals with DS is associated with more severe illness following viral infections, such as the viral infection during the H1N1 influenza pandemic ^4^, respiratory syncytial virus (RSV) ^5^, and may also lead to worse outcomes due to coronavirus disease 2019 (COVID-19). The cytokine storm reported to be associated with COVID-19 ^6^, refers to an excessive immune response to external stimuli and is thought to be a predictor of increased severity of SARS-CoV-2 infection and of poor COVID-19 prognosis ^7^. Admission to intensive care units (ICU) following COVID-19 diagnoses was found to be associated with higher plasma levels of many cytokines ^7^. Individuals with DS frequently display elevated levels of cytokines in blood, even in the absence of viral infection, and a proinflammatory profile that includes increased natural killer (NK) cells, CD8, decreased CD19, and an increased spontaneous production of interferon gamma, TNFα, and IL-10 ^8,9^. Recent findings in this fast-evolving field, however, suggest COVID-19 may not be characterized by a cytokine storm after all ^10^. Moreover, it is also reasonable to speculate that increased levels of cytokines, particularly interferon gamma, may actually help patients fight off the SARS-CoV-2 infection.

DS is associated with several co-occurring health conditions including obesity, diabetes, hypotonia, obstructive sleep apnea, craniofacial dysmorphogenesis, and congenital heart defects, as well as gastroesophageal reflux that may contribute to the increased risk of respiratory tract infection ^1,11–13^. Moreover, adults with DS have many age-related health conditions including Alzheimer’s disease ^14^, that often occur 20-25 years earlier than in individuals without DS which is a profile that suggests premature aging ^1,11,14^. Yet, there is also evidence that persons with DS are protected from cardiovascular disease, including hypertension ^15^. Additionally, the incidence and severity of various comorbid conditions are quite variable among people with DS. Therefore, it is difficult to predict whether individuals with DS are particularly susceptible to COVID-19 and its complications.

In the US and many European countries, individuals with DS older than 30 years of age tend to live in group homes and other assisted-living situations. Furthermore, it is not uncommon for those with DS in their 50 and 60’s to dwell in nursing homes for prolonged periods. These living spaces, which many times are occupied by several individuals and frequently visited by multiple support staff, have been the sites of several well-documented COVID-19 outbreaks in older individuals in the general population ^16^. Therefore, one issue of particular importance is ascertaining whether individuals with DS are at increased risk for hospitalization and mortality following COVID-19 diagnosis at a younger age than their counterparts without DS. However, up to now, most reports of COVID-19 in individuals with DS have been based on less than 10 individuals ^17,18^. One recent report used primary care data from the UK, but included a small sample of individuals with DS who have died (n = 27), and may be unrepresentative of mild cases, due to patients not having access to their primary care physicians during the peak of the pandemic ^19^.

In order to obtain large scale information on specific vulnerabilities, clinical presentation, and outcomes of COVID-19 in individuals with DS, the Trisomy 21 Research Society (T21RS) launched an online survey, with input from stakeholders including leading clinicians and DS organisations. The survey was designed to address three primary questions: 1) What are the presenting signs and symptoms of COVID-19 in individuals with DS and are these different than in individuals without DS? 2) Are those with DS at an increased risk for complications associated with SARS-CoV-2 infection? 3) Is the profile of risk factors associated with poor outcomes for COVID-19 in persons with DS the same as in the general population? Here we report on the data collected from Europe, United States (US), Latin America, India, as well as hospital data from other sources.

## METHODS

### T21RS DS survey

An online survey was developed in March 2020, to identify cases of COVID-19 among individuals with DS (a copy of the survey is provided in the supplementary material). Two equivalent surveys were developed, one to be completed by caregivers/family members of COVID-19 affected individuals with DS and one to be completed by clinicians. Most often, either the caregiver or the clinician completed the survey per person. When both forms were completed for the same individual, information was combined to maximize the amount of available data per person. The survey collected information on: 1) basic demographics, 2) living situation during the pandemic, 3) pre-existing health conditions, vaccinations and medications, 4) SARS-CoV-2 testing, 5) presenting signs and symptoms, 6) hospital and ICU admission, 7) complications associated with SARS-CoV-2 infection (clinician only), 8) medications and treatments used during COVID-19 illness (clinician only), and 9) status at last evaluation. The survey was implemented through REDCap ^20,21^, a survey and database management system, and was hosted at Emory University. As we include data from the early phase of the COVID-19 pandemic and the testing capacities differ between countries, we defined cases as individuals with DS of all ages, who tested positive for SARS-CoV-2 or reported signs or symptoms of COVID-19. The survey was disseminated through clinical routes (e.g. Down syndrome medical interest group listservs and health service providers), Down syndrome associations in the US, India, Spain, UK, France, Italy, Germany, Brazil, and Spanish-speaking Latin America, and Down syndrome registries (e.g., NIH DS-Connect®) as well as via the T21RS website. Each institution that planned to disseminate the survey within health services obtained IRB/ethics approval (Table S1). The study was performed according to the Declaration of Helsinki and national guidelines and regulations for data privacy. All data were anonymized according to good clinical practice guidelines and data protection regulations. For this analysis, we included data from all cases that were entered between April 9, 2020 and October 22, 2020.

### Statistical analysis

Only individuals with information on age and gender were included in the analyses. We used descriptive statistics to show the demographic information, COVID-19 signs, symptoms, outcomes, and comorbidities of the participants included in our analyses. To prevent study participants from being included twice in the analyses (reported by both caregiver and clinician), we excluded duplicated participants based on age, gender and country and other specific demographics.

To contrast the COVID-19 related signs, symptoms, and medical complications with those observed in the general population, we compared the T21RS DS cases to data from the UK ISARIC4C survey. The ISARIC4C survey is a prospective observational cohort study engaging 208 acute-care hospitals in England, Wales, and Scotland ^22^. In this study, we included 59,025 hospitalized patients with COVID-19 from the ISARIC4C survey, which were entered between February 2020 and July 9, 2020 (downloaded on July 24, 2020). Among the 59,025 hospitalized patients, 109 individuals had DS (58,916 did not). Of these, 100 had complete data on age, gender and ethnicity. These 100 individuals were matched to 400 individuals without DS (controls) from the same survey (matching 1:4) as well as to 100 hospitalized individuals with DS from the T21RS survey (matching 1:1) based on age, gender and ethnicity (see supplementary tables S2 and S3 for characteristics of the matched samples). Fisher’s exact test was used to probe for differences in the prevalence of signs, symptoms, and medical complications between matched individuals with DS (from the ISARIC4C and T21RS surveys) and without DS (controls from the ISARIC4C survey).

To compare the mortality rates of hospitalized COVID-19 patients with and without DS in different age groups, we combined data from different sources. Case counts and mortality rates among hospitalized patients from the general population were estimated from individuals without DS included in the UK ISARIC4C survey combined with published hospital reports from Spain ^23^ and New York City ^24^. Mortality rates among hospitalized patients with DS were based on combined data from the UK ISARIC4C survey and hospitalized individuals from the T21RS survey. In addition, we used the matched samples described above to perform a comparison of mortality rates corrected for age, gender and ethnicity using logistic regression. To test whether differences in mortality rates of individuals with and without DS can be explained by differences in the prevalence of known risk factors for COVID-19-related mortality, we adjusted the association analyses for chronic cardiac disease, chronic pulmonary disease, chronic kidney disease, liver disease, obesity, chronic neurological disorder, dementia, malignant neoplasm ^22^. In a sensitivity analysis, we analysed the association between DS and COVID-19-related mortality in the whole ISARIC4C sample (52,142 individuals without DS and no missing data for age, gender and ethnicity, 100 individuals with DS) adjusted for i) age, gender and ethnicity and ii) additionally for chronic cardiac disease, chronic pulmonary disease, chronic kidney disease, liver disease, obesity, chronic neurological disorder, dementia, malignant neoplasm. In addition, we conducted age-stratified association analyses for individuals younger than 40 years versus 40 years or older.

To identify risk factors associated with adverse outcomes of COVID-19 in individuals with DS, we conducted adjusted logistic regression analyses in symptomatic COVID-19 cases from the T21RS survey data. Associations between potential risk factors and admission to hospital or all-cause mortality were analysed. Associations with age and gender were adjusted for the data source (caregiver versus clinician survey) and associations with living situation, level of intellectual disability, and comorbidities were adjusted for age, gender, data source and country of residence. We only included comorbidities that were present in at least 15% of our study samples to reduce the burden of multiple testing and the chance of false positives due to small numbers of cases.

We did all data analyses using R (version 4.0.0).

### Role of the funding sources

Funding sources provided help with dissemination of the online surveys and partial support of the biostatisician’s efforts. They were not involved in: the study design; collection, analysis, or interpretation of data; writing of the manuscript; or in the decision to submit the paper for publication.

## RESULTS

### Description of study population

Of the 1906 records in the T21RS database entered between April 9, 2020 and October 22, 2020, 1103 individuals had signs and symptoms of COVID-19 or a positive test result and provided information on age and gender. After removing potential duplicates, the final sample size was 1046 COVID-19 patients with DS (Figure S1). The majority of these cases were symptomatic (981 (94.5%)) and 861 (83%) were tested for COVID-19 (750 (91%) with a positive test result) (Table S4). Of the 1046 COVID-19 patients with DS, 591 (57%) were reported by a clinician and 455 (43%) by a family member or caregiver (Table 1). The largest number of cases were from India (405 (40%)), followed by the United States (163 (16%)), Spain (155 (15%)) and Brazil (75 (7%)). The mean age was 29 years (SD=18); cases reported by a clinician were on average 15 years older than cases reported by a family member or caregiver. The majority of cases lived at home with their family (712 (73%)). The proportion of cases living in a residential care facility was higher in the clinician than family/caregiver reports (134 (26%) versus 23 (5%)). The majority of cases (784 (93%)) had full trisomy 21 and moderate level of intellectual disability (580 (62%)), similar to that observed in the population of individuals with DS at large ^11^. About half of the cases were admitted to hospital (581 (56%)), of which 279 (50%) were admitted to an ICU and 207 (29%) were connected to a mechanical ventilator. More than half of the cases (547 (55%)) had recovered from COVID-19 at the last evaluation and 131 individuals (13%) had died.

**Table 1.**
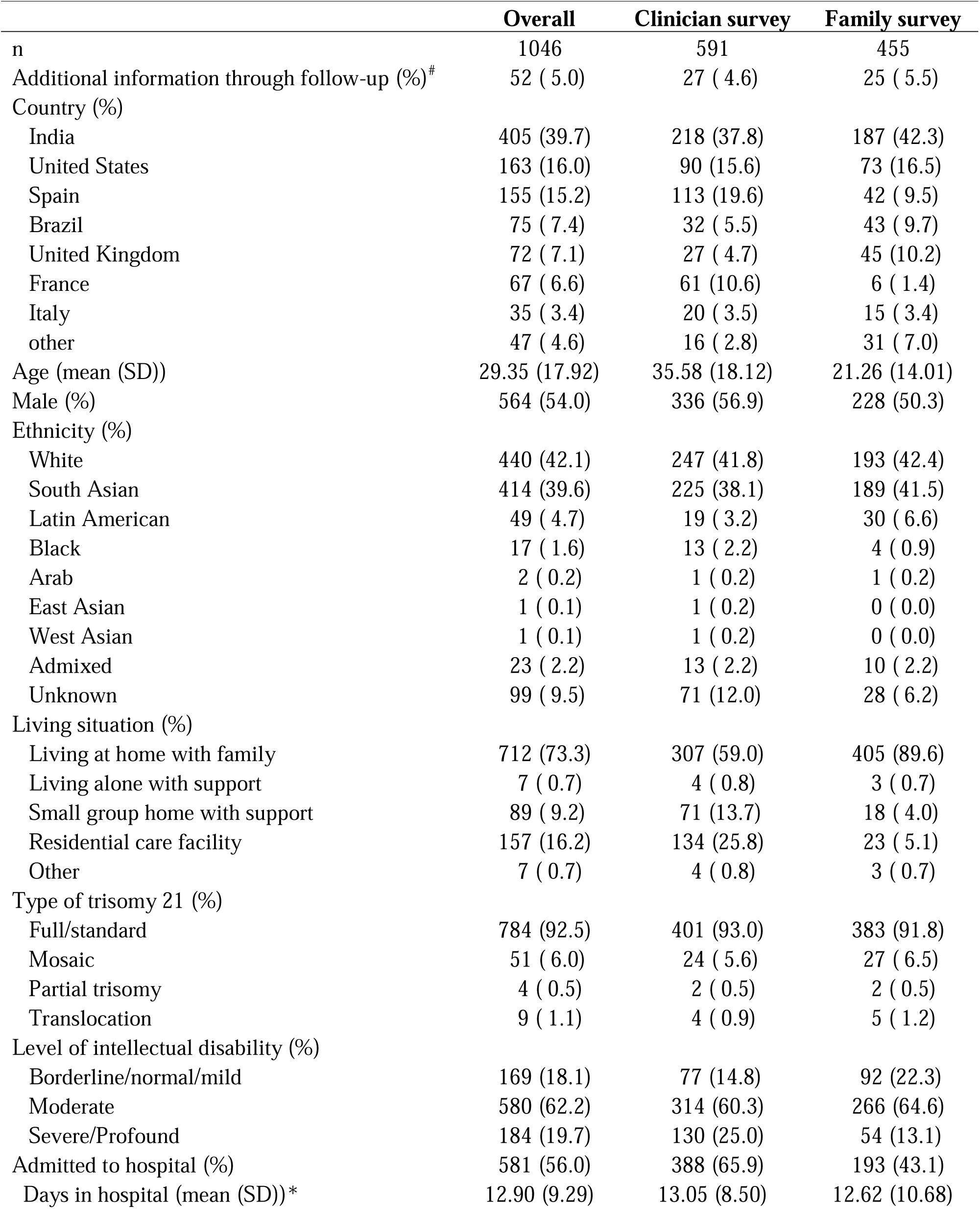
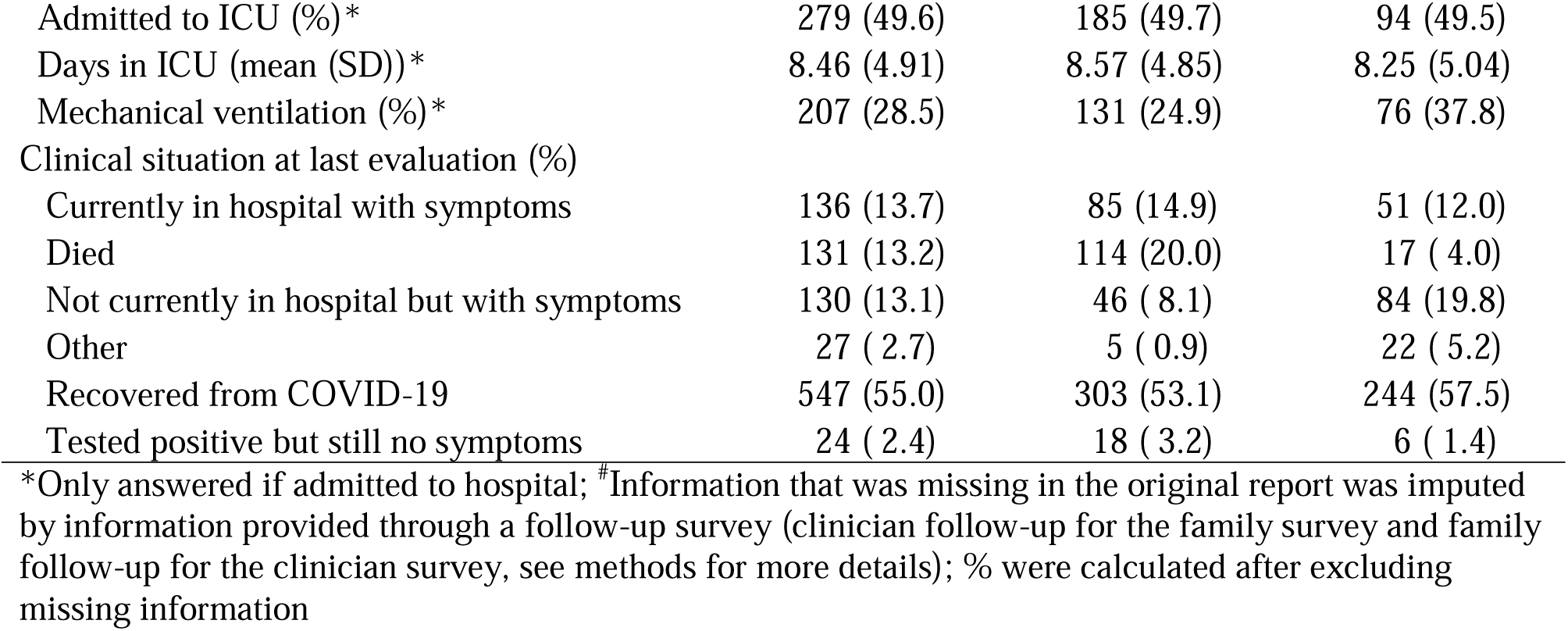
**T21RS** study characteristics grouped by data source (reported by clinician versus family/caregiver)

### Signs and symptoms presenting in COVID-19 patients with DS and without DS

The leading signs and symptoms related to COVID-19 in individuals with DS were fever, cough and shortness of breath (Figure 1, Table 2), which is in line with those reported in the general population (Table 2, ISARIC4C controls). Nasal signs (excess nasal drainage or ‘runny nose’) and pharyngitis (‘sore throat’) were also common among COVID-19 patients from the T21RS survey, particularly in children and adolescents (younger than 20 years), where nasal signs were present in 229 (61%) and sore throat in 165 children and adolescents (44%) independent of the data source (family or clinician survey, Figure S2). Chills (33%), abdominal pain (32%), nausea or vomiting (26%) and muscle or joint pain (18%) were particularly common in the age group 20-30 years. Signs and symptoms that were more prevalent in individuals with DS 40 years and older include extreme fatigue or confusion (30%) and not eating or drinking (21%).

**Table 2.** Signs and symptoms related to COVID-19 in individuals with and without Down syndrome. Hospitalized individuals with Down syndrome from the UK ISARIC4C and the T21RS surveys are compared to matched individuals without Down syndrome (controls) from the ISARIC4C survey. The 100 individuals with Down syndrome reported through the UK ISARIC4C survey were matched to 400 individuals without Down syndrome (controls) from the same survey (matching 1:4) as well as to 100 individuals with Down syndrome from the T21RS survey (matching 1:1). The matching was based on age, gender and ethnicity (see supplementary tables S2 and S3 for characteristics of the matched samples).

|  | ISARIC4C controls |  | ISARIC4C individuals with Down syndrome |  |  | T21RS matched individuals with Down syndrome |  |  |
| --- | --- | --- | --- | --- | --- | --- | --- | --- |
|  | n (%) | N | n (%) | N | p value<br>(comparison with controls) | n (%) | N | p value<br>(comparison with controls) |
| Cough<sup>1</sup> | 270 (71.8) | 376 | 65 (67.7) | 96 | 0.451 | 54 (54.0) | 100 | **0.001** |
| Fever | 294 (76.4) | 385 | 64 (68.1) | 94 | 0.112 | 80 (80.0) | 100 | 0.505 |
| Sore throat | 39 (13.0) | 301 | 8 (12.1) | 66 | 1.000 | 17 (17.0) | 100 | 0.320 |
| Runny nose<sup>2</sup> | 13 (4.4) | 293 | 2 (3.3) | 61 | 1.000 | 16 (16.0) | 100 | **<0.001** |
| Joint pain or muscle aches | 90 (28.8) | 313 | 3 (4.8) | 63 | **<0.001** | 11 (11.0) | 100 | **<0.001** |
| Fatigue/Malaise<sup>3</sup> | 149 (46.4) | 321 | 33 (45.8) | 72 | 1.000 | 32 (32.0) | 100 | **0.011** |
| Shortness of breath | 248 (68.9) | 360 | 66 (74.2) | 89 | 0.368 | 76 (76.0) | 100 | 0.176 |
| Disturbance or loss of taste/smell<sup>4</sup> | 15 (9.3) | 162 | 0 (0.0) | 27 | 0.206 | 3 (3.0) | 100 | 0.076 |
| Headache | 55 (18.3) | 301 | 2 (3.1) | 64 | **0.001** | 14 (14.0) | 100 | 0.362 |
| Altered consciousness or confusion<sup>3</sup> | 61 (17.6) | 346 | 25 (32.9) | 76 | **0.004** | 32 (32.0) | 100 | **0.003** |
| Abdominal pain | 60 (18.2) | 330 | 10 (14.3) | 70 | 0.493 | 11 (11.0) | 100 | 0.094 |
| Vomiting/nausea | 92 (27.0) | 341 | 8 (10.7) | 75 | **0.003** | 14 (14.0) | 100 | **0.008** |
| Diarrhoea | 77 (23.1) | 334 | 10 (12.3) | 81 | **0.034** | 17 (17.0) | 100 | 0.216 |
| Skin rash or skin ulcers<sup>5</sup> | 13 (4.0) | 321 | 4 (5.1) | 78 | 0.754 | 2 (x) | 44 | 0.700 |

<sup>1</sup>ISARIC4C: combination of the signs “cough”, “cough with sputum production” and “cough bloody sputum / haemoptysis”; <sup>2</sup> T21RS survey: this sign is referred to “nasal signs” rather than runny nose; <sup>3</sup>In the T21RS survey, this is referred to as “extreme fatigue, confusion, difficulty staying alert”; <sup>4</sup>ISARIC4C: combined ageusia and anosmia; <sup>5</sup>ISARIC4C: combined skin rash and skin ulcers; n/a: these questions were not included in the T21RS survey; % were calculated after excluding missing information.

**Figure 1.**
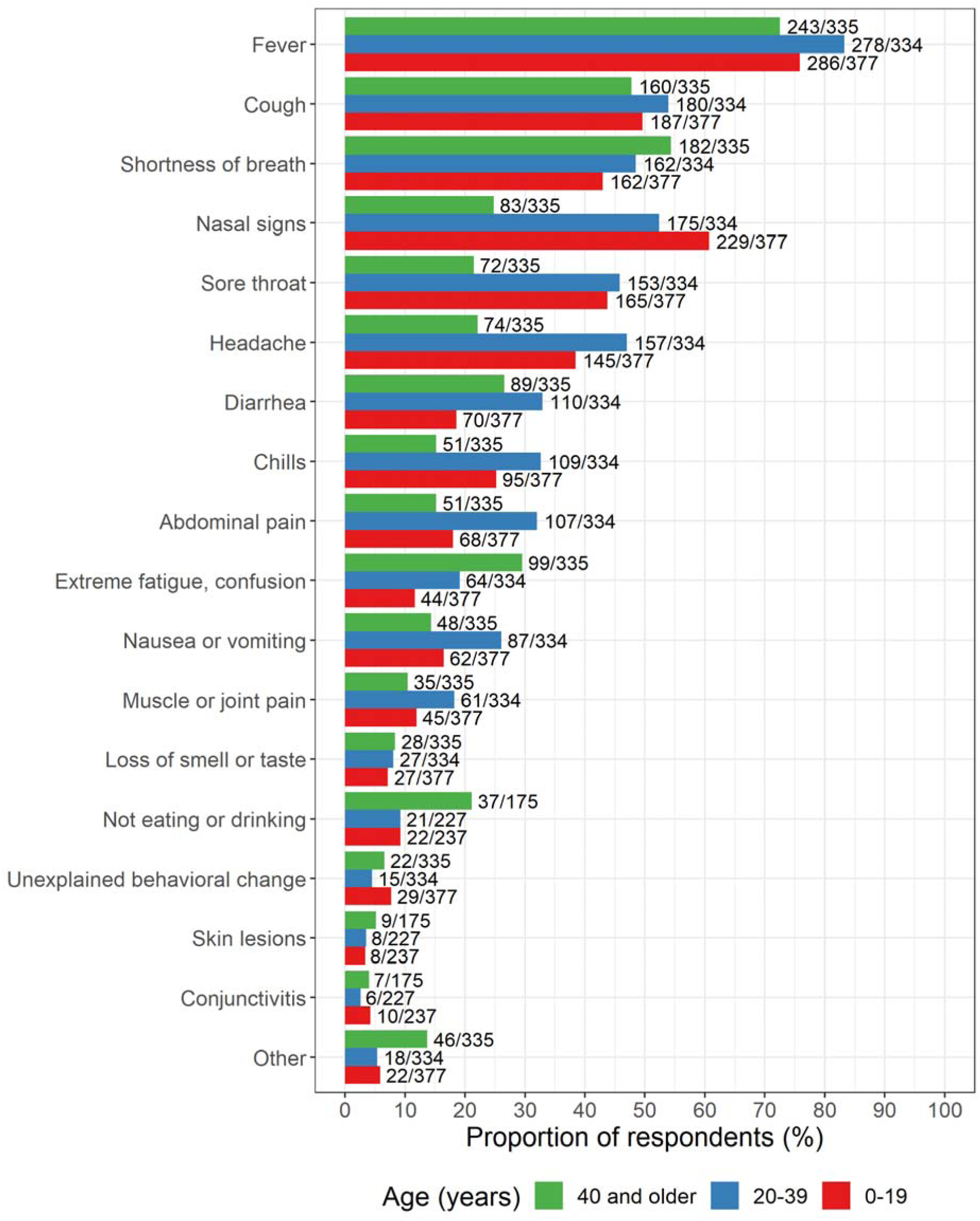
Signs and symptoms reported among the COVID-19 cases with Down syndrome grouped by age (T21RS survey). The signs and symptoms “not eating or drinking”, “conjunctivitis” and “skin lesions” were added in the second wave of the survey (smaller sample size for these symptoms).

Family members and caregivers tended to report more signs and symptoms, and more often reported extreme fatigue and confusion, than clinicians (Figure S2). The signs and symptoms reported for cases with a positive test result did not differ from those reported for the whole study sample (Figure S3). Furthermore, reported signs and symptoms were similar between individuals with mild-to-moderate and severe intellectual disability (Figure S4). Individuals admitted to hospital had more signs and symptoms in general compared with those who were not hospitalized, with shortness of breath being particularly more common among individuals admitted to hospital (Figure S5).

Comparing the signs and symptoms of hospitalized individuals with DS to matched individuals without DS (based on age, gender, and ethnicity) showed that joint pain or muscle aches was less frequently reported in individuals with DS (controls: 29%, ISARIC4C DS: 5%, (p < 0.001), T21RS DS: 11% (p<0.001), Table 2). Altered consciousness or confusion was more frequently reported in individuals with DS (controls: 18%, ISARIC4C DS: 33% (p = 0.004), T21RS DS: 32% (p =0.003)) and vomiting/nausea was less often reported in individuals with DS (controls: 27%, ISARIC4C DS: 11% (p = 0.003), T21RS DS: 14% (p = 0.008)). Nasal signs were more often reported in individuals with DS only from the T21RS survey, which might be due to differences in study design (ISARIC4C: hospital reports by clinicians; T21RS: voluntary online survey by family or clinicians).

### Medical complications among COVID-19 patients with and without DS

Overall, 360 (60%) of the T21RS DS cases reported by clinicians had developed medical complications due to COVID-19 (medical complications were not included in the caregiver survey, Table S5). Experiencing medical complications was correlated with higher mortality rates. The most prevalent complications were viral pneumonia (36%) and acute respiratory distress syndrome (34%), followed by secondary bacterial pneumonia (17%) and septic shock (11%). Of the individuals who died, 69% had viral pneumonia and 85% an acute respiratory distress syndrome. The prevalence of medical complications increased by age. While 64 (41%) of the 0-19 year-olds developed medical complications due to COVID-19, the numbers increased to 103 (65%) in 20-39 year-olds and 193 (69%) in individuals with DS 40 years or older (Table S6).

Comparing the medical complications observed in hospitalized patients with DS to matched patients without DS (based on age, gender and ethnicity) showed that pulmonary complications (pneumonia and acute respiratory syndrome) were more common in individuals with DS (Table 3). These differences were significant for all three pulmonary complications comparing the T21RS data to the ISARIC4C controls (p < 0.01) and suggestive for the comparison of viral pneumonia within the ISARIC4C data (p = 0.059). There was no difference in the frequency of cardiac complications, anaemia, or acute renal injury. Among those with COVID-19-related viral pneumonia, individuals with DS were more likely to die than individuals without DS (controls: 23%, ISARIC4C DS: 53% (p < 0.001); T21RS DS: 57% (p < 0.001)). A similar trend was observed for acute respiratory syndrome and cardiac complications, but the differences were only significant for the comparison between the T21RS data to the ISARIC4C controls. Overall, medical complications were more common in the T21RS data than in individuals with DS from the ISARIC4C survey and the frequencies differed between countries, with India, Spain and United States reporting the highest numbers of medical complications (Table S7). Thus, observed differences might be due to differences in study design or due to small numbers (e.g., cardiac complications).

**Table 3.** Medical complications related to COVID-19. Prevalence of cardiac complications and other medical complications that occurred in at least 10% of the matched control group and the associated mortality rate. Hospitalized individuals with Down syndrome from the UK ISARIC4C and the T21RS surveys are compared to matched individuals without Down syndrome (controls) from the ISARIC4C survey. The 100 individuals with Down syndrome reported through the UK ISARIC4C survey were matched to 400 individuals without Down syndrome (controls) from the same survey (matching 1:4) as well as to 100 individuals with Down syndrome from the T21RS survey (matching 1:1). The matching was based on age, gender and ethnicity (see supplementary tables S2 and S3 for characteristics of the matched samples).

| **A. Prevalence of medical complications among hospitalized patients** |  |  |  |  |  |  |  |  |
| --- | --- | --- | --- | --- | --- | --- | --- | --- |
|  | **ISARIC4C controls** |  | **ISARIC4C individuals with Down syndrome** |  |  | **T21RS individuals with Down syndrome** |  |  |
|  | n (%) | N | n (%) | N | p value<br>(comparison with controls) | n (%) | N | p value<br>(comparison with controls) |
| Viral pneumonia<sup>1</sup> | 151 (42.7) | 354 | 49 (53.8) | 91 | 0.059 | 60 (83.3) | 72 | <0.001 |
| Bacterial pneumonia | 35 (10.0) | 351 | 14 (15.2) | 92 | 0.189 | 8 (32.0) | 25 | 0.004 |
| Acute Respiratory Syndrome | 47 (13.0) | 362 | 17 (17.9) | 95 | 0.245 | 41 (60.3) | 68 | <0.001 |
| Cardiac complications<sup>2</sup> | 32 (8.6) | 370 | 6 (6.3) | 95 | 0.535 | 5 (8.5) | 59 | 1.000 |
| Anaemia | 43 (11.7) | 367 | 11 (12.0) | 92 | 1.000 | n/a | n/a | n/a |
| Acute renal injury/Acute renal failure | 51 (13.8) | 369 | 12 (12.8) | 94 | 0.867 | 11 (17.7) | 62 | 0.434 |
| **B. Associated mortality rate** |  |  |  |  |  |  |  |  |
| Viral pneumonia<sup>1</sup> | 34 (22.5) | 151 | 26 (53.1) | 49 | <0.001 | 34 (56.7) | 60 | <0.001 |
| Bacterial pneumonia | 11 (31.4) | 35 | 6 (42.9) | 14 | 0.516 | 2 (25.0) | 8 | 1.000 |
| Acute Respiratory Syndrome | 17 (36.2) | 47 | 8 (47.1) | 17 | 0.563 | 31 (75.6) | 41 | <0.001 |
| Cardiac complications<sup>2</sup> | 10 (31.3) | 32 | 2 (33.3) | 6 | 1.000 | 5 (100.0) | 5 | 0.007 |
| Anaemia | 14 (32.6) | 43 | 4 (36.4) | 11 | 1.000 | n/a | n/a | n/a |
| Acute renal injury/Acute renal failure | 19 (37.3) | 51 | 5 (41.7) | 12 | 1.000 | 7 (63.6) | 11 | 0.177 |

<sup>1</sup>T21RS survey: In the 1<sup>st</sup> wave of our survey, we only asked for pneumonia without differentiating between viral and bacterial pneumonia. As most clinicians answered this question with information on COVID-19-related viral pneumonia, the information provided on “pneumonia” in the 1<sup>st</sup> wave of the survey was combined with “viral pneumonia” for the analyses. Combination of cryptogenic organizing pneumonia (COP), pneumothorax, pleural effusion, bronchiolitis;
<sup>2</sup>ISARIC4C: Combination of congestive heart failure, endocarditis / myocarditis pericarditis, myocarditis / pericarditis, cardiomyopathy, cardiac arrhythmia, cardiac ischemia; % were calculated after excluding missing information.

### Mortality rates among COVID-19 patients with and without DS

Overall, 13% of the 1046 COVID-19 patients with DS from the T21RS DS data died (Table 1). Eighty-seven percent of the fatal outcomes were reported by clinicians; 13% were reported by family members or caregivers. The mean age of cases who died was 51 years (SD=12), whereas the mean age of cases who did not die was 27 years (SD=17) (Table S8).

Among individuals with DS who were hospitalized with COVID-19, we observed an increased mortality rate from age 40, while in the general population mortality rate increased at age 60 (Figure 2A, Tables S9 and S10). Under the age of 40, 17/250 (7%) of hospitalized COVID-19 patients with DS died, compared to 139/4815 (3%) in the general population (Figure 2A).

**Figure 2.**
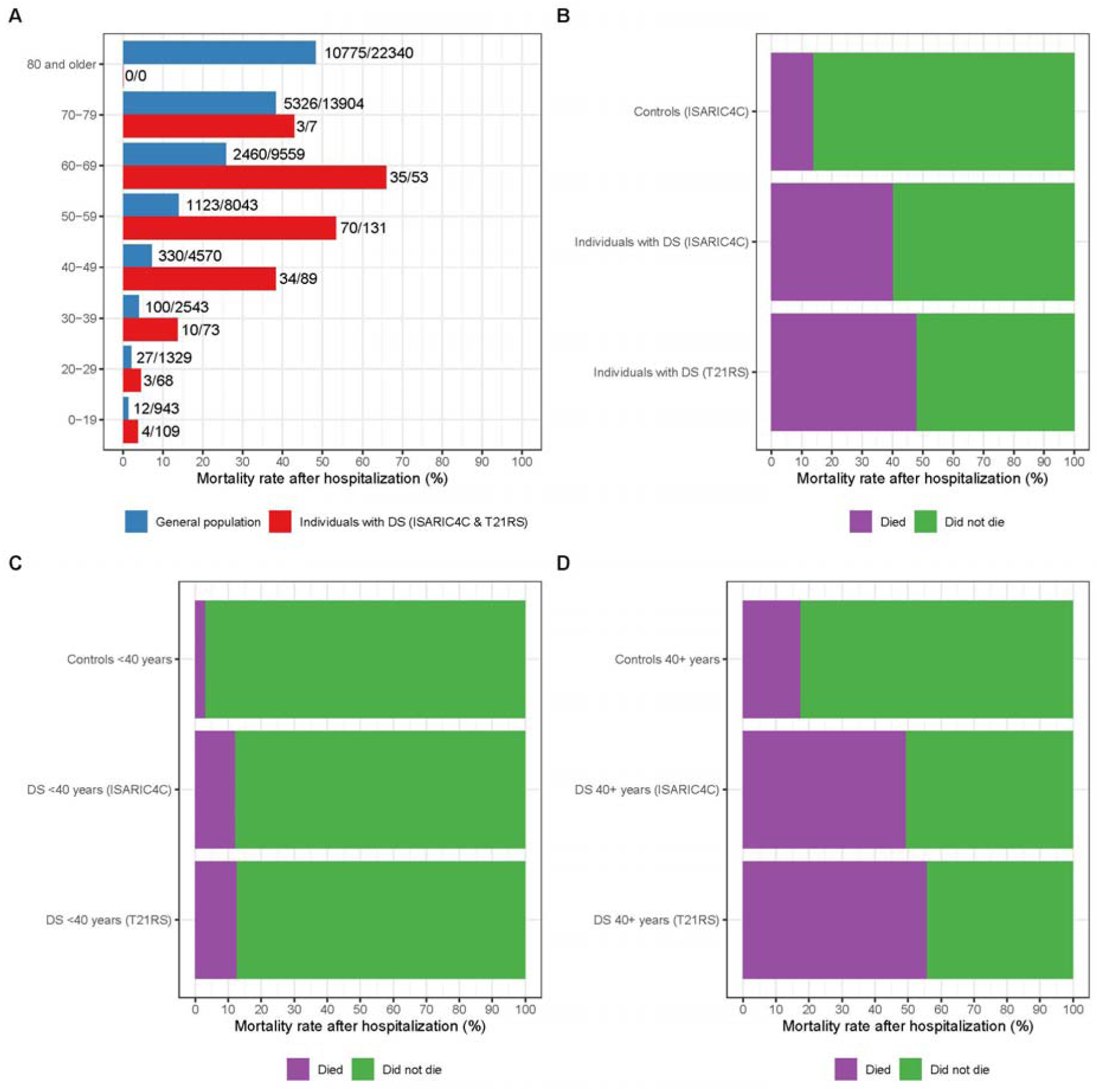
Mortality rates among patients hospitalized with COVID-19. **A**. Age distribution of the proportion of deaths among individuals with DS (combined data from the T21RS and ISARIC4C surveys), who were hospitalized with COVID-19, in comparison to hospitalized cases of COVID-19 from the general population (combined data from the UK ISARIC4C survey (patients without DS), NYC ^24^ and Spain ^23^). **B**., **C. and D**. Mortality rates among hospitalized individuals. Hospitalized individuals with Down syndrome from the UK ISARIC4C and the T21RS surveys are compared to matched individuals without Down syndrome (controls) from the ISARIC4C survey. The 100 individuals with Down syndrome reported through the UK ISARIC4C survey were matched to 400 individuals without Down syndrome (controls) from the same survey (matching 1:4) as well as to 100 individuals with Down syndrome from the T21RS survey (matching 1:1). As samples were matched based on age, gender and ethnicity, mortality rates are corrected for these characteristics (see supplementary tables S2 and S3 for more information on the matched samples). We had information on the outcome of disease in 91/100 individuals from the matched T21RS dataset. **B**. All matched individuals. **C**. Matched individuals younger than 40 years of age. **D**. Matched individuals 40 years of age or older.

Comparing mortality rates among the matched ISARIC4C and T21RS samples, individuals with DS hospitalized with COVID-19 were around three times more likely to die than individuals without DS, assuming the same age, gender and ethnicity (Figure 2B, individuals with and without DS from ISARIC4C: risk ratio (RR) = 2.9 (95%-CI = 2.1; 3.8), individuals with DS from T21RS vs. individuals without DS from ISARIC4C: RR = 3.5 (95%-CI = 2.6; 4.4)). Adjusting for known COVID-19 related risk factors for mortality (chronic cardiac disease, chronic pulmonary disease, chronic kidney disease, liver disease, obesity, chronic neurological disorder, dementia, malignant neoplasm), reduced the RR to 2.5 (95%-CI = 1.5; 3.7) (ISARIC4C DS versus controls, Table S11A). That means that even after accounting for known risk factors, individuals with DS were still 2.5 times more likely to die after being hospitalized with COVID-19. However, mortality rates of individuals younger than 40 years of age were substantially lower (3% in controls, 12% in matched individuals with DS from the ISARIC4C survey and 13% in matched individuals from the T21RS survey), which is in line with mortality rates in individuals under the age of 40 with and without DS from the whole study sample (Figure 2A, Tables S5 and S6). The differences in mortality rates between these younger individuals with and without DS were not significant after adjusting for known risk factors for COVID-19 mortality (chronic cardiac disease, chronic pulmonary disease, chronic kidney disease, liver disease, obesity, chronic neurological disorder) (RR=2.4, 95%-CI = 0.1; 12.9, Table S11B), suggesting that excess mortality in younger individuals with DS was potentially due to differences in comorbid conditions or to small sample sizes of those who died. However, comparing individuals with DS younger than 40 and those over 40 admitted to hospital while adjusting for comorbidities known to be associated with COVID-19 and those common in DS, suggest that the reduction in risk for mortality is between 68% (95%-CI 18 - 93%; ISARIC4C dataset) and 92% (95%-CI 84 – 96%; T21RS dataset) lower in the younger individuals compared with those over 40 (table S12).

### Risk Factors for hospitalization and mortality among COVID-19 patients with DS

Age was the strongest risk factor for hospitalization and mortality in individuals with DS from the T21RS survey (Figure 3, Table S13). Every additional 10 years of age increased the odds for hospitalization by a factor of 1.75 (95%-CI = 1.47; 2.07) and the odds for mortality by a factor of 2.41 (95%-CI = 2.02; 2.89). We further observed increased odds for hospitalization for males (OR (95%-CI) = 1.51 (1.16; 1.97)), individuals with obesity (OR (95%-CI) = 2.03 (1.44; 2.87)), diabetes (OR (95%-CI) = 1.93 (1.2; 3.1)) and individuals with a congenital heart defect (OR (95%-CI) = 1.46 (1.05; 2.03)). We observed increased odds for mortality for males (1.75, 95% CI = 1.16-2.63) and individuals with Alzheimer’s disease/dementia (OR (95%-CI) = 2.13 (1.1; 4.12)). Other risk factors were not significant.

**Figure 3.**
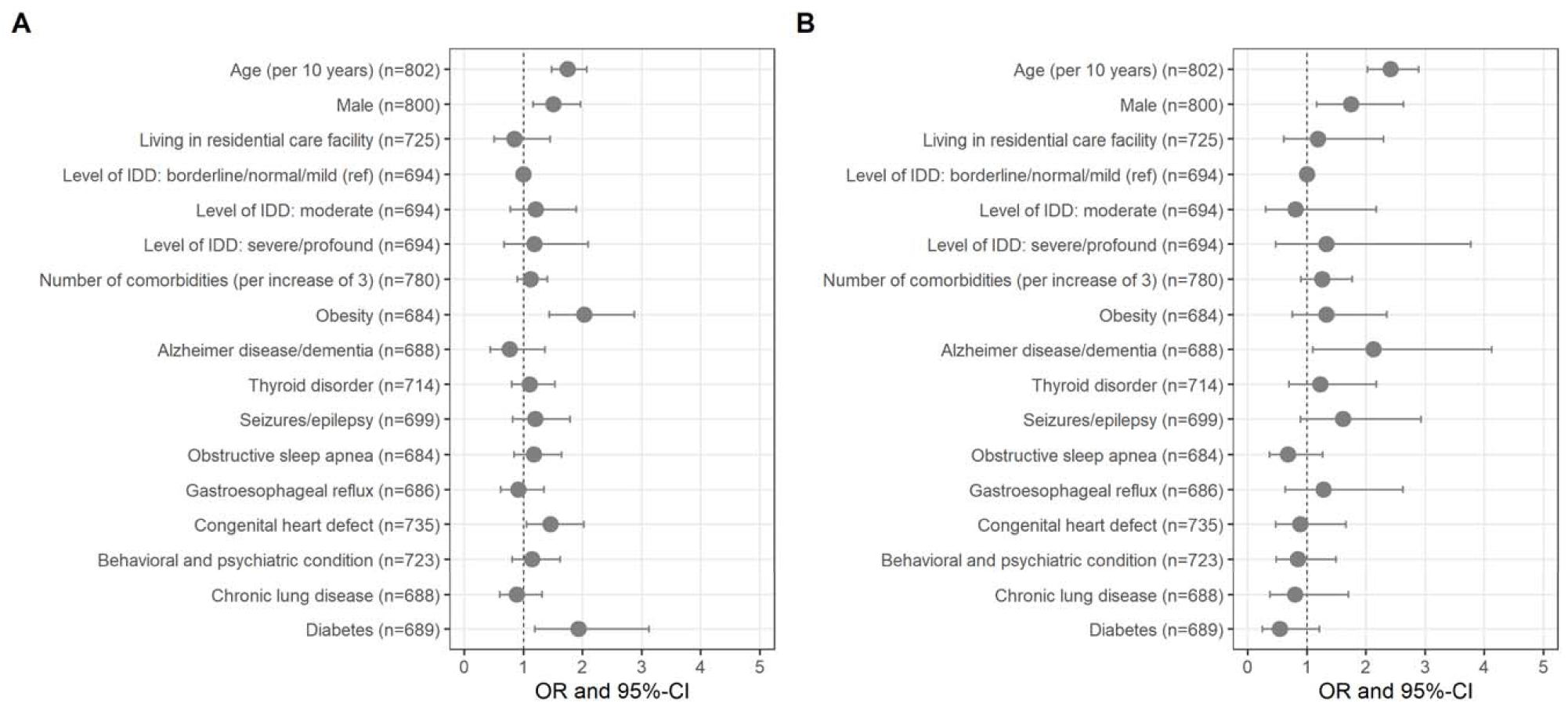
Risk factors associated with adverse outcomes of COVID-19 in individuals with Down syndrome. Associations with **A**. hospitalization and **B**. mortality in symptomatic COVID-19 patients with Down syndrome from the T21RS survey estimated in adjusted logistic regression models (odds ratios (OR) and 95%-confidence intervals (95%-CI)). Associations with age and gender were adjusted for the data source (caregiver versus clinician survey) and associations with living situation, level of IDD and comorbidities were adjusted for age, gender, data source and country of residence. Abbreviations: IDD, intellectual and developmental disabilities; ref, reference category.

## Discussion

DS is associated with specific co-occurring conditions and an immune-response dysfunction that may impact the risk of individuals with this disorder for having a more severe course of illness following infection by SARS-CoV-2. To examine this question, we conducted an international online survey to collect COVID-19 related information on signs and symptoms, course of disease, and potential risk factors in more than 1,000 patients with COVID-19 and DS. Our analyses showed that while the leading signs and symptoms related to COVID-19 are similar to those in the general population, patients with DS who are hospitalized with COVID-19 were more likely to experience a severe course of disease with higher rates of medical complications and mortality than the general population, particularly adults older than 40 years of age.

Similar to the general population, fever, cough and shortness of breath were the leading signs and symptoms related to COVID-19 in individuals with DS ^22^. From the general population, we know that COVID-19 is usually a mild disease in children and often accompanied with a few upper respiratory symptoms, including nasal congestion and runny nose ^25^. Interestingly, we found that nasal signs were much more common in individuals from our T21RS survey than in individuals with and without DS from the ISARIC4C survey (hospital reports from the UK), which may relate to earlier signs of COVID-19 occurring prior to hospitalization or to different reporting by families/caregivers. This highlights the importance of including data from community-dwelling individuals with COVID-19 in addition to routine hospital reports as hospital reports are less likely to capture mild signs and symptoms that might be important for early detection of disease.

Altered consciousness or confusion were more commonly observed in individuals with DS than in the general population. As this sign could be an indicator of disease severity, it suggests that individuals with DS are more likely to become severely ill with COVID-19 than individuals without DS, or that they might be more severely ill at the time of hospitalization due to delayed diagnosis. Joint pain or muscle aches and vomiting/nausea were less commonly reported in individuals with vs. without DS. There is little evidence that people with DS experience both acute and chronic pain with a lower frequency than the rest of the population ^26^. However, there is some evidence that parental perception of pain is less discriminant for children with DS than for their siblings without DS ^27^. Therefore, it may be difficult to detect, discriminate, and report pain in patients with DS. Like pain, nausea has to be self-reported by the patient. Hence, it is possible that both pain and nausea were underreported in individuals with DS and poorly detected and documented by clinicians. This possibility highlights the importance of including family members/caregivers in the clinical assessment of COVID-19 patients with DS to avoid underrepresentation of self-reported symptoms.

Mortality rates could only be compared among hospitalized COVID-19 patients with and without DS. We know that mortality rates among all individuals with COVID-19 can be sensitive to reporting bias and to the amount of testing that is done in the population. For the US, the CDC reported a hospitalization rate of 14% (16% of these were admitted to an ICU) and a mortality rate of 5% ^28^. In the T21RS survey sample, the admission rate to the ICU was 50% among all hospitalized individuals with DS; this proportion was almost identical between clinician (49.7%) and caregiver reports (49.5%) and is much higher than the one reported in the general population in the UK (17%) ^22^. This can be another indicator of more severe outcomes of disease in individuals with DS.

Hospitalized individuals with DS were around three times more likely to die than individuals without DS, assuming the same age, gender and ethnicity, and this association was only slightly attenuated when adjusted for known risk factors for COVID-19-related mortality. However, additional analysis showed that children and young adults (younger than age 40) were about 90% less likely to die once admitted to hospital with COVID-19 in comparison to older adults with DS. Nevertheless, pulmonary medical complications (pneumonia and Acute Respiratory Syndrome) were more prevalent and deadlier in individuals with DS, which is in line with the more severe complications during other viral respiratory infections such as influenza and RSV ^5^.

Age was the strongest risk factor for hospitalization and mortality in COVID-19 patients with DS, which is in line with findings from the general population ^22^. Importantly, we observed an increased mortality rate from age 40, at much earlier age than in the general population. This observation is consistent with the reduced median lifespan of individuals with DS, which is estimated to be between 55 to 60 years ^6^, i.e., around 20 years younger than that of the general population. Many markers of accelerated aging have been well documented in individuals with DS ^6^, chief among them being the premature development of Alzheimer’s disease pathology and dementia. This emphasizes the need to protect older individuals with DS from SARS-Cov-2 infections. While mortality rates among children and young adults with DS were low, they were still higher than in individuals without DS from the same age groups; however more data are needed to confirm this pattern.

Additional risk factors for hospitalization were male gender, obesity, diabetes and congential heart defect and those for mortality were male gender and Alzheimer’s disease/dementia. As most of these are known risk factors for severe outcomes of COVID-19 ^22^, our findings suggest that the risk factors for individuals with DS are similar to those observed in the general population. However as obesity, dementia and congenital heart defect occur at higher rate in DS than the general population, their role in increasing risk for severe outcomes in this population is highly significant.

Except for congenital heart defects, we did not identify any other risk factors that were specific for individuals with DS. More studies with larger sample sizes may be needed to investigate whether other common comorbidities of DS (e.g. obstructive sleep apnea, gastroesophageal reflux, or seizures) increase the risk for a severe course of COVID-19. Of note, we did not find any indication that living in residential care facilities or the level of intellectual disability increases the risk for COVID-19 related hospitalization or mortality. It has been reported that individuals living in residential care facilities are more vulnerable to COVID-19 infection ^29^, but it has also been noted by other authors “that people with [intellectual and developmental disabilities] living in congregate care settings can benefit from a coordinated approach to infection control, case identification and cohorting” ^30^. Possibly as evidence of adequate management or rapid response in many of these facilities, we observed in our study population that infected individuals in congregate care settings were not more vulnerable to adverse outcomes of disease than those living with their families.

The T21RS survey is by far the largest assessment of individuals with DS who were infected with SARS-CoV-2 that has been reported to date. Including both family and clinician reports has the advantage of capturing information from a broad spectrum of severity of COVID-19 and individuals from all age groups, which is in contrast to most previous large-scale studies on COVID-19 that are based on hospital reports^22–24^. In addition, our survey has been distributed in many different countries, which increased the sample size and added diversity to our study. Nonetheless, as a sample of convenience, our study population might not be representative of the population of individuals with DS at large.

The primary limitation of this study is that not all of the COVID-19 cases were confirmed by COVID-19-specific laboratory tests. Given that our survey was launched at the beginning of the pandemic when testing was scarce, we would have lost more than 20% of our cases (particularly from Spain, France, and Italy) if we were to include only individuals with laboratory-confirmed COVID-19. Nonetheless, the comparison with individuals from the general population was restricted to patients hospitalized with COVID-19, providing an added level of confidence for the clinical diagnosis even when laboratory tests were not reported. Another limitation of our study is the inclusion of cases from different countries and thus different health systems. Some countries had more COVID-19 cases than others, health care systems differ substantially between countries (e.g. India versus Western European countries) and some countries experienced a surge in cases later than others, so that they had more time to prepare and develop treatment strategies. To address this potential bias, we conducted the comparison to the general population for cases that were matched based on age, gender and ethnicity; and adjusted the association analyses with risk factors for hospitalization and mortality for country of residence.

The comparison with data from the general population also has some limitations. The inclusion criteria are different between studies due to limited testing capacity ^22–24^. Furthermore, we could only compare data from hospitalized individuals and it is unknown whether COVID-19 patients with DS are admitted more or less often to hospitals and which factors or reasons may affect their rate of hospitalization. Lastly, the samples for the matched comparison were from the UK, whereas the T21RS study samples came from many different countries and country-specific differences could have biased our results. Furthermore, we cannot determine whether the 11 matched hospitalised T21RS cases from the UK were also part of the ISARIC4C survey.

There are several open questions we could not answer with our study. As our study only included individuals infected with SARS-CoV-2, we could not address whether individuals with DS are more likely to get infected. Furthermore, due to our limited sample size, we were not able to identify any risk factors for a severe course of COVID-19 that might be specific for individuals with DS. Another question we could not specifically address is whether COVID-19 patients with DS are subject to significant discrimination that might have affected the offer of invasive interventions. The high rates of ICU admissions within our study sample, however, do not support this hypothesis. Yet, it is possible that patients with DS may have been admitted to ICU later than patients without DS due to delay in diagnosis.

Further research is required to establish whether treatments are equally effective in individuals with DS, and whether specific treatments are required to reduce the increased rate of severe outcomes.^1^ There should be a focus on preventing or ameliorating severe outcomes, which includes prioritisation for immunisation (influenza, pneumococcus, and, when available, SARS-Cov-2). In addition, our study sample only includes a small number of asymptomatic cases (6%), likely due to lack of routine testing; thus, it remains unknown whether trisomy 21 or DS-related comorbidities alter the risk of having symptoms after infection with SARS-CoV-2.

Our results have implications for preventive and clinical management of individuals with COVID-19 and DS. In terms of sheltering/shielding individuals to prevent infection, more caution should be taken with individuals over age 40, which is about 20 years younger than the typical at-risk group in the general population. Individuals with DS aged 40 and older have a three-fold increased risk for severe outcomes compared to the general population, which is not explained by additional comorbidities. Bearing in mind limited data, the present study also suggests that younger individuals are unlikely to develop severe disease, but more data are needed to confirm this supposition. The T21RS survey shows that clinicians and caregivers should primarily look for the same signs and symptoms seen in the general population (e.g. fever, cough, shortness of breath) when monitoring for COVID-19 in individuals with DS. Milder signs and symptoms such as a ‘runny nose’ can also be a sign of COVID-19, particularly in younger patients with DS, which is often underreported. With this information in mind, it is also safe to assume that most of the recommendations given to the general population, such as limiting exposure by social distancing and access to safe activities with respiratory protection, are also important to prevent SARS-CoV-2 infection and spread of COVID-19 among those with DS. Additional strategies include prioritising individuals with DS for immunisations, including for flu and pneumococcus, to reduce the risk for severe outcomes following infection with SARS-CoV-2.

## Supporting information

Author group (T21RS COVID-19 Initiative)

Caregiver/family survey

Clinician survey

## Data Availability

The data code books for the caregiver/family and clinician T21RS survey are provided in Supplementary Materials.
The REDCap data dictionary can be provided on request to the corresponding author.
Written requests for the use of the anonymized data collected through the T21RS survey will be reviewed for approval by the T21RS COVID-19 Initiative.

## Acknowledgments

Trisomy 21 Research Society (T21RS) COVID-19 Taskforce developed the survey, with the financial and dissemination support of Down Syndrome Affiliates in Action (DSAIA), Down Syndrome Medical Interest Group-USA (DSMIG-USA), GiGi’s Playhouse, Jerome Lejeune Foundation, LuMind IDSC Foundation, Matthews Foundation, National Down Syndrome Society (NDSS), and the National Task Group on Intellectual Disabilities and Dementia Practices (NTG). These and other international Down syndrome organizations are members of the T21RS COVID-19 stakeholders advisory group that provided advice to inform the design of the survey questions and interpretation of results, including the Global Down Syndrome Foundation (USA), DSA (UK), DSMIG (UK), DSMIG (USA), DSRF-UK, DSi, DSE international, Trisomie21-France, Down España, National Down Syndrome Congress (NDSC), Down Madrid, Fundació Catalana Síndrome de Down (Spain), EDSA, Royal College of Psychiatrists, CoorDown (Italy), Associazione Italiana Persone Down (AIPD; Italy), AFRT (France), Fundación Iberoamericana Down 21 (Spain), FIADOWN (Latin America), Federação Brasileira das Associações de Síndrome de Down (Brazil) and the European Down Syndrome Association. We acknowledge the contribution of DS-Connect® (The Down Syndrome Registry) which is supported by the *Eunice Kennedy Shriver* National Institute of Child Health and Human Development (NICHD), NIH for the dissemination of the T21RS survey. We also wish to thank the many families and clinicians who contributed to the survey.

This report is independent research which used data provided by the MRC funded ISARIC 4C Consortium and which the Consortium collected under a research contract funded by the National Institute for Health Research. The views expressed in this publication are those of the author(s) and not necessarily those of the ISARIC 4C consortium.

AH is supported by the HERCULES Center (NIEHS P30ES019776) and the LuMind IDSC Foundation. The REDCap survey and database management system at Emory University was supported by Library Information Technology Services grant support (UL1 TR000424). ACSC is supported by the Alana USA Foundation, Awakening Angels Foundation, and the Infectious Diseases Society of America (IDSA). MD is supported by the Centre for Genomic Regulation Severo Ochoa excellence grant, the CIBER of Rare Diseases, DURSI 2017SGR595, and acknowledges the Spanish Ministry of Science, Innovation and Universities (MSIU) to the EMBL partnership, the Centro de Excelencia Severo Ochoa and CERCA (GenCat). AS is supported by the MRC (MR/S011277/1; MR/S005145/1; MR/R024901/1), Lumind IDSC, The LeJeune Foundation and the European Commission (H2020 SC1 Gene overdosage and comorbidities during the early lifetime in Down Syndrome GO-DS21-848077). ML was supported by the National Institute for Health Research (NIHR) Biomedical Research Centre based at UCL Great Ormond Street Institute of Child Health/Great Ormond Street Hospital NHS Foundation Trust). The Research Programme on Biomedical Informatics (GRIB) is a member of the Spanish National Bioinformatics Institute (INB), funded by ISCIII and EDER (PT17/0009/0014). The DCEXS is a ‘Unidad de Excelencia María de Maeztu’, funded by the AEI (CEX2018-000782-M). The GRIB is also supported by the Agència de Gestió d’Ajuts Universitaris i de Recerca (AGAUR), Generalitat de Catalunya (2017 SGR 00519).

This study was also supported by the Fondo de Investigaciones Sanitario (FIS), Instituto de Salud Carlos III (PI18/00335 to MCI, PI14/01126 and PI17/01019 to JF), partly jointly funded by Fondo Europeo de Desarrollo Regional, Unión Europea, Una manera de hacer Europa; the Jérôme Lejeune Foundation (No.1319 Cycle 2019B to MCI); the National Institutes of Health (NIA grants 1R01AG056850 - 01A1; R21AG056974 and R01AG061566 to JF); Departament de Salut de la Generalitat de Catalunya, Pla Estratègic de Recerca i Innovació en Salut (SLT006/17/00119 to JF); and Fundació La Marató de TV3 (20141210 to JF).

## Declaration of interests

The authors have nothing to declare.

## Contributions

AH: Planned and conducted the statistical analyses; the major contributor in writing the manuscript. ACSC: Major contributor to design and distribution of the survey; led the efforts in Brazil and involved in translation; involved in interpretation of findings; major contributions to the writing of the manuscript. MD: Major contributor to design and distribution of the survey; involved in interpretation of findings; led the Spanish Taskforce. RAB: Contributed to statistical analyses, including analysis of ISARIC4C dataset. SB: Contributed to design of the work, acquisition of data. NTB: Contributed to design of the work, acquisition of data. ACB: Led the survey in Brazil; translated the questionnaire into Portuguese; obtained official approval for the study in Brazil; contribution to the design of some questions in the survey. AC: Contributed to design of the work, acquisition of data. MCI: Contributed to the design of the work, acquisition of data. BAC: Participated in study design/modification; data collection; discussion of data analysis. SG: Led and organized Indian survey on DS; coordinated among the researchers, clinicians and DS families; obtained ethical clearance for the survey in India. ML: Contributed to design of survey; interpretation of findings. CM: Helped with Ethics approval in Spain; survey translation into Spanish; design of protocol; distribution of survey; data entry. MAM: Contributed to writing the report for the Ethical Committee in Spain; involved in the design of the survey, translating it into Spanish. MCO: Recruitment of cases in Spain and contribution to the design of the psychiatric questions in the survey. DRdA: Data collection, interpretation of results. ASR: Design of the survey, data acquisition, collection and interpretation, translation in French; local ethics submission. LAR: Data management for the T21RS survey; data analyses. GS: Contributed to design of the work; acquisition of data. DV: Led the survey in Italy; obtained official approval for the study in Italy; contribution to the design of some questions in the survey. SLS: Contributed to the design and development of the survey; data verification, analyses and interpretation, major contributions to the writing of the manuscript. AS: Involved in conception and design of the T21RS survey and ISARIC4C analysis; major contributions to the writing of the manuscript as well as UK PI. All authors critically reviewed early and final versions of the manuscript.

## Data Sharing

- The data code books for the caregiver/family and clinician T21RS survey are provided in Supplementary Materials.
- The REDCap data dictionary can be provided on request to the corresponding author.
- Written requests for the use of the anonymized data collected through the T21RS survey will be reviewed for approval by the T21RS COVID-19 Initiative.

## Notes

### Competing Interest Statement

The authors have declared no competing interest.

### Author Declarations

Each institution that planned to disseminate the survey within health services obtained IRB/ethics approval (Table S1).

