## Supplementary Material for "An international survey on the impact of COVID-19 in individuals with Down syndrome"

*SG: Cytogenetics and Genomics Reserach Unit. Department of Zoology, University of Calcutta.Kolkata. West Bengal, India*

*ML: UCL- Great Ormond Street Institute of Child Health, London, United Kingdom; Whttington NHS Trust, London, United Kingdom; Down Syndrome Medical Interest Group, London, United Kingdom*

*CM: DOWN ESPAÑA, Barcelona, Spain*

*MAM: Research Programme on Biomedical Informatics (GRIB), Hospital del Mar Medical Research Institute and DCEXS Universitat Pompeu Fabra, Barcelona, Spain*

*MO: Department of Psychiatry, Research Institute i+12. Hospital Universitario 12 de Octubre. Madrid, Spain*

*DR: Department of Internal Medicine and Instituto de Investigación Biomédica-La Princesa, Hospital Universitario de La Princesa, Madrid, Spain*

*ASR: Institut Jérôme Lejeune, Paris, France*

*LR: Department of Epidemiology, Rollins School of Public Health, Emory University, Atlanta, Georgia, USA*

*GS: Department of Developmental Neuroscience, IRCCS Fondazione Stella Maris, Pisa, Italy;* *Department of Clinical and Experimental Medicine, University of Pisa, Pisa, ItalyDV: Pediatric Unit, Bambino Gesù Children's Hospital, IRCCS, Rome, Italy*

*SLS:* *Department of Human Genetics, School of Medicine, Emory University, Atlanta, Georgia, USA*

*AS: Institute of Psychiatry, Psychology, and Neuroscience, Department of Forensic and Neurodevelopmental Sciences, King’s College London, London, United Kingdom; The London Down Syndrome (LonDownS) Consortium, London, United Kingdom; South London and the Maudsley NHS Foundation Trust*

**Corresponding author:**

Andre Strydom, MD

*Institute of Psychiatry, Psychology, and Neuroscience, Department of Forensic and Neurodevelopmental Sciences, King’s College London, London, United Kingdom; The London Down Syndrome (LonDownS) Consortium, London, United Kingdom; South London and the Maudsley NHS Foundation Trust*

[+44 20 7848 5701](tel:+44%2020%207848%205701)

| **Table S1.** Each institution that planned to disseminate the survey obtained IRB/ethics approval. | |
| --- | --- |
| **Spain** | The study was approved by the Hospital del Mar ethics committee (CEIC Parc de Salut Mar, CEim 2020/9197) |
| **United Kingdom** | The T21RS survey was approved by the Health research agency (HRA) 20/HRA/2452. The ISARIC4C analysis was approved by the study board (IDAMAC) |
| **Brazil** | This study was approved by the Brazilian Federal Ethics Committee (CONEP, CAAE: 30847520.8.0000.0071) |
| **Emory University, U.S.A.** | This study was deemed exempt from human subjects research under 45 CFR 46.104(d)(2i) (IRB ID: STUDY00000386) |
| **Italy** | The study was approved by the Bambino Gesù children’s Hospital Ethics Committee (2091_OPBG_2020) |
| **Advocate Health Care Institutional Review Board** | Determined to have Exempt Status IRB# 20-151ET |
| **France** | This study was approved by CPP Sud Mediterranée IV dated 29/04/2020 (ID RCB 2020-A00940-39) |
| **Ludwig-Maximilians-Universität (LMU), Munich, Germany** | Determined to have Exempt Status (IRB ID: 20-573 KB) |
| **India** | The study was approved by the Ethics Committee constituted by the University of Calcutta (CU/BIOETHICS/HUMAN/2306/3044/2020) |

| **Table S2. Characteristics of matched samples.** Hospitalized individuals with Down syndrome from the UK ISARIC4C and the T21RS surveys are compared to matched individuals without Down syndrome (controls) from the ISARIC4C survey. The 100 individuals with Down syndrome reported through the UK ISARIC4C survey were matched to 400 individuals without Down syndrome (controls) from the same survey (matching 1:4) as well as to 100 individuals with Down syndrome from the T21RS survey (matching 1:1). The matching was based on age, gender and ethnicity. | | | |
| --- | --- | --- | --- |
|  | **ISARIC4C Controls** | **ISARIC4C individuals**  **with Down syndrome** | **T21RS matched individuals**  **with Down syndrome^1^** |
| *Age group*  < 20  20 – 29  30 – 39  40 – 49  50 – 59  60 – 69  70 - 79 | 36  24  40  68  128  84  20 | 9  6  10  17  32  21  5 | 9  6  10  17  32  25  1 |
| *Gender*  Male  Female | 208  192 | 52  48 | 52  48 |
| *Ethnicity*  Black  South Asian  White  Other | 20  16  324  40 | 5  4  81  10 | 4  8  81  7 |
| ^1^Due to the limited sample size in the age group 70-79 years in the T21RS sample, 4 individuals from this age group were matched to individuals from the age group 60-69 years. We tried to match individuals on age, gender and ethnicity. If we were not able to find a match, we matched individuals only on age and gender. | | | |

| **Table S3.** Study characteristics of the whole T21RS sample in comparison to the subset of individuals matched with the individuals with Down syndrome from the ISARIC4C survey. | | |
| --- | --- | --- |
|  | **Overall** | **Matched individuals**  **for comparison with**  **ISARIC4C survey** |
| n | 1046 | 100 |
| Additional information through follow-up (%)^#^ | 52 ( 5.0) | 2 (2.0) |
| Country (%) |  |  |
| India | 405 (39.7) | 8 (8.1) |
| United States | 163 (16.0) | 25 (25.3) |
| Spain | 155 (15.2) | 29 (29.3) |
| United Kingdom | 75 ( 7.4) | 11 (11.1) |
| France | 72 ( 7.1) | 0 (0) |
| Brazil | 67 ( 6.6) | 6 (6.1) |
| Italy | 35 ( 3.4) | 15 (15.2) |
| other | 47 ( 4.6) | 5 (5.1) |
| Age (mean (SD)) | 29.35 (17.92) | 47.33 (16.5) |
| Male (%) | 564 (54.0) | 52 (52.0) |
| Ethnicity (%) |  |  |
| White | 440 (42.1) | 81 (81.0) |
| South Asian | 414 (39.6) | 8 (8.0) |
| Latin American | 49 ( 4.7) |  |
| Black | 17 ( 1.6) | 4 (4.0) |
| Arab | 2 ( 0.2) |  |
| East Asian | 1 ( 0.1) |  |
| West Asian | 1 ( 0.1) |  |
| Admixed | 23 ( 2.2) |  |
| Unknown | 99 ( 9.5) | 7 (7.0) |
| Living situation (%) |  |  |
| Living at home with family | 712 (73.3) | 38 (42.2) |
| Living alone with support | 7 ( 0.7) | 1 (1.1) |
| Small group home with support | 89 ( 9.2) | 13 (14.4) |
| Residential care facility | 157 (16.2) | 38 (42.2) |
| Other | 7 ( 0.7) |  |
| Type of trisomy 21 (%) |  |  |
| Full/standard | 784 (92.5) | 56 (98.2) |
| Mosaic | 51 ( 6.0) | 1 (1.8) |
| Partial trisomy | 4 ( 0.5) |  |
| Translocation | 9 ( 1.1) |  |
| Level of intellectual disability (%) |  |  |
| Borderline/normal/mild | 169 (18.1) | 10 (12.2) |
| Moderate | 580 (62.2) | 54 (65.9) |
| Severe/Profound | 184 (19.7) | 18 (22.0) |
| Admitted to hospital (%) | 581 (56.0) | 100 (100.0) |
| Days in hospital (mean (SD))* | 12.90 (9.29) | 10.63 (7.69) |
| Admitted to ICU (%)* | 279 (49.6) | 26 (28.3) |
| Days in ICU (mean (SD))* | 8.46 (4.91) | 9.86 (5.78) |
| Mechanical ventilation (%)* | 207 (28.5) | 18 (20.9) |
| Clinical situation at last evaluation (%) |  |  |
| Currently in hospital with symptoms | 136 (13.7) | 9 (9.3) |
| Died | 131 (13.2) | 42 (43.3) |
| Not currently in hospital but with symptoms | 130 (13.1) | 5 (5.2) |
| Other | 27 ( 2.7) | 2 (2.1) |
| Recovered from COVID-19 | 547 (55.0) | 39 (40.2) |
| Tested positive but still no symptoms | 24 ( 2.4) |  |
| *Only answered if admitted to hospital; ^#^Information that was missing in the original report was imputed by information provided through a follow-up survey (clinician follow-up for the family survey and family follow-up for the clinician survey, see methods for more details) | | |

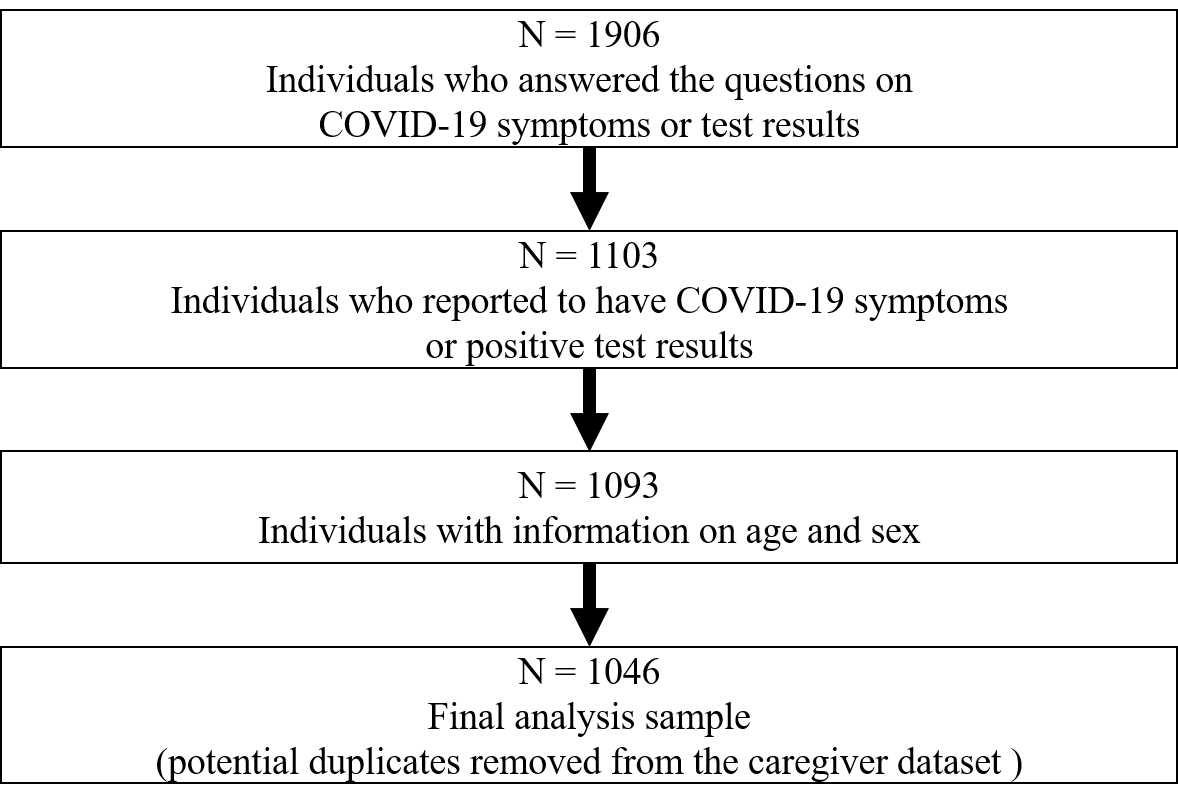

**Figure S1. Flow chart of the T21RS survey samples.**

| **Table S4.** Proportion of the T21RS analysis sample with COVID-19 symptoms or positive test results. | |
| --- | --- |
|  | **Overall** |
| n | 1046 |
| Signs and symptoms of COVID-19 (%) | 981 (94.5) |
| COVID-19 testing performed (%) | 861 (83.0) |
| Reason for COVID-10 testing (%) |  |
| Contact with affected individuals | 102 (12.0) |
| Routine testing | 21 ( 2.5) |
| Symptoms were present | 729 (85.6) |
| Results of the testing (%) |  |
| Negative | 58 ( 7.0) |
| Pending | 16 ( 1.9) |
| Positive | 750 (91.0) |

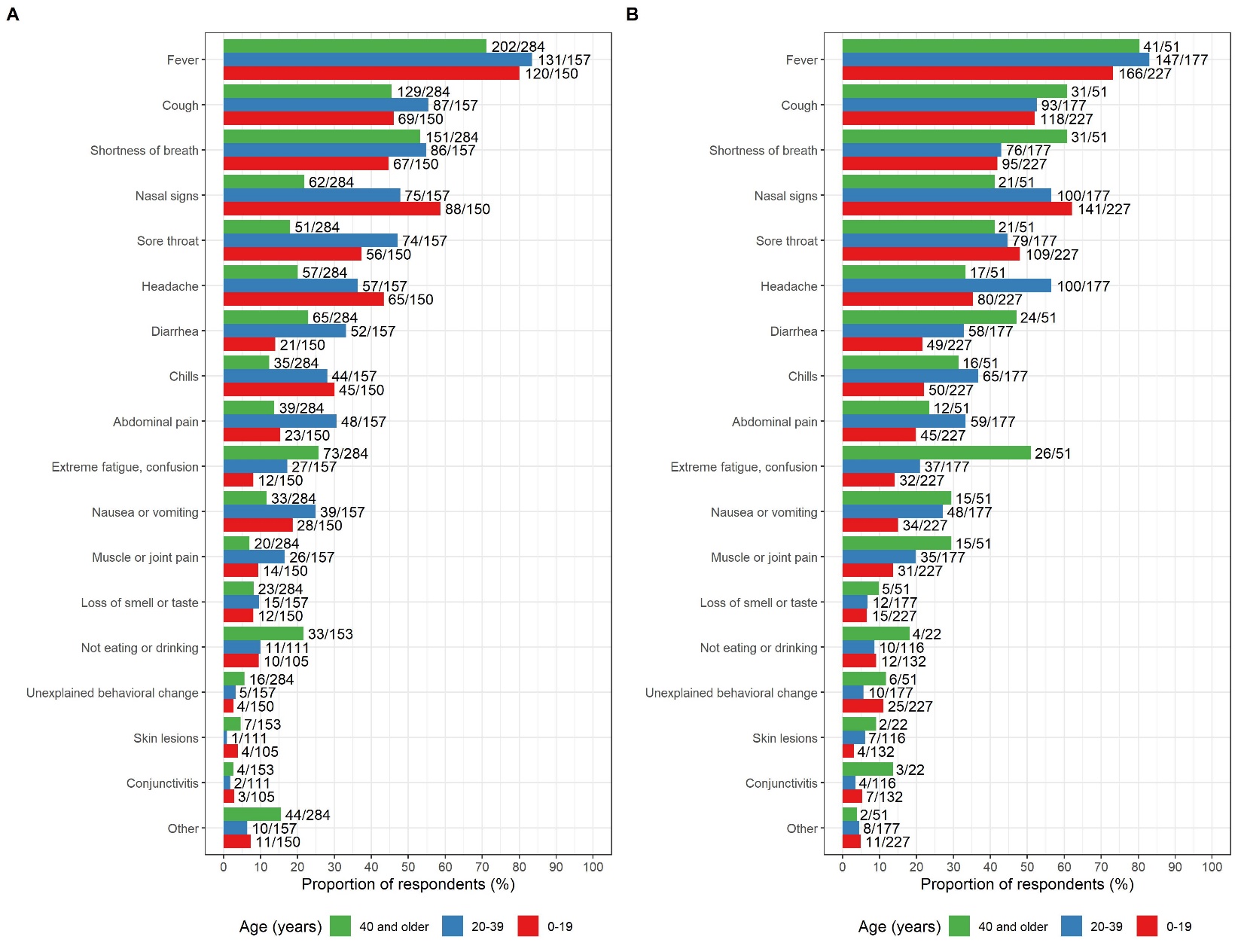

**Figure S2. Signs and symptoms reported among the COVID-19 cases with Down syndrome (T21RS survey) grouped by age and stratified by: A. Clinician reports, B. Family member/caregiver reports.** The symptoms “not eating or drinking”, “conjunctivitis” and “skin lesions” were added in the second wave of the survey (smaller sample size for these symptoms).

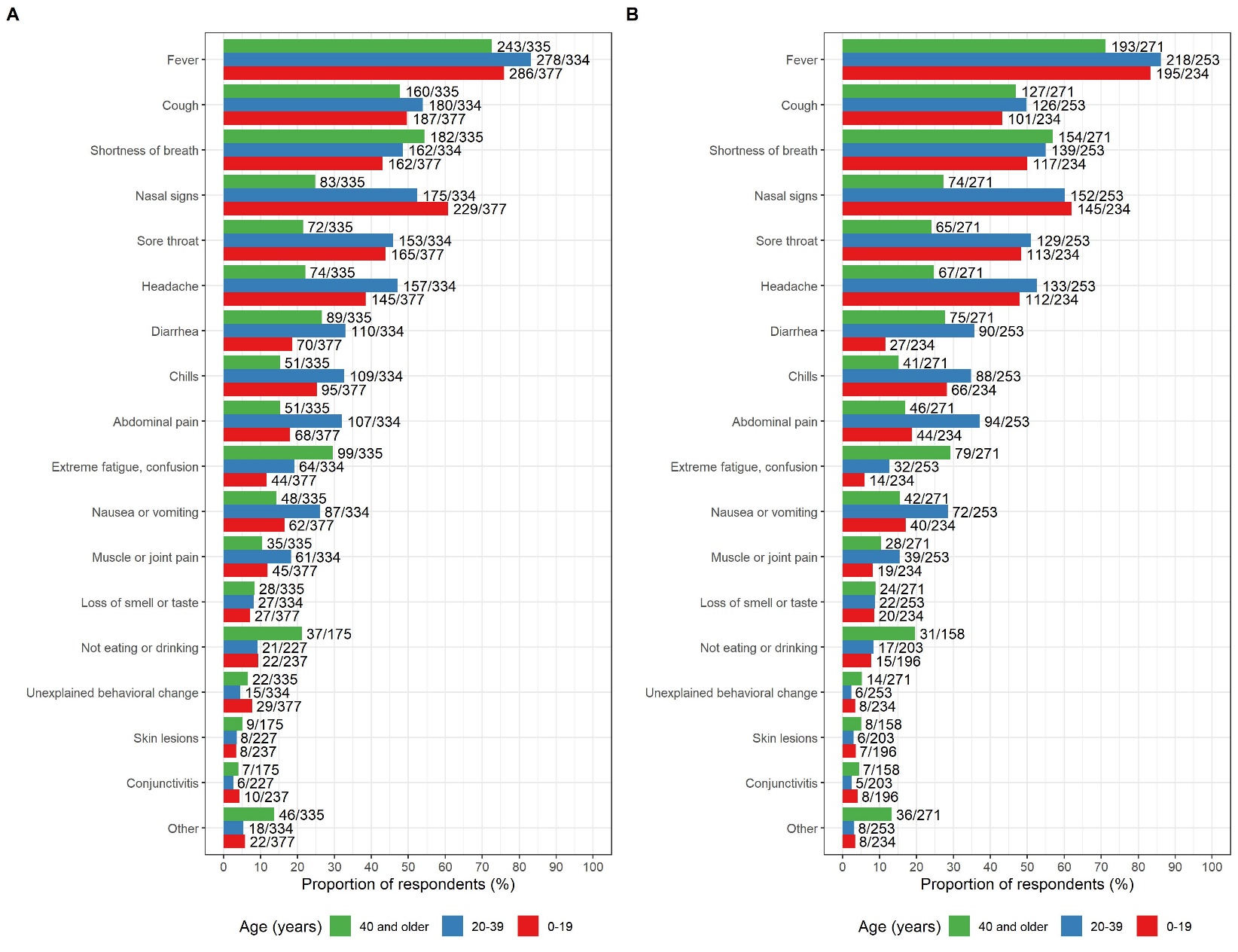

**Figure S3.** **Signs and symptoms reported among the COVID-19 cases with Down syndrome (T21RS survey) grouped by age and stratified by: A. All cases, B. Cases with positive test result.** The symptoms “not eating or drinking”, “conjunctivitis” and “skin lesions” were added in the second wave of the survey (smaller sample size for these symptoms).

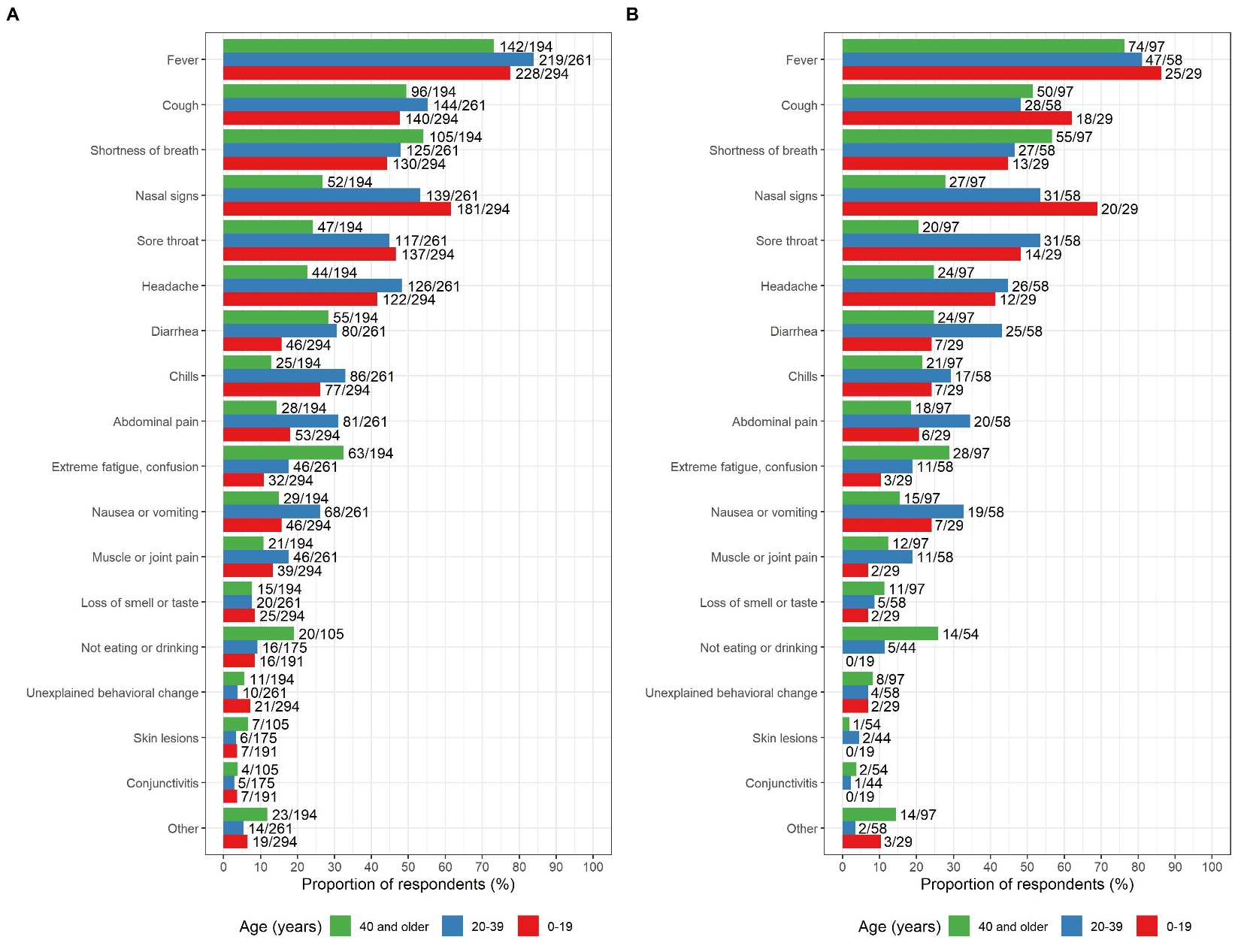

**Figure S4. Signs and symptoms reported among the COVID-19 cases with Down syndrome (T21RS survey) grouped by age and stratified by level of intellectual disability**: **A. mild to moderate, B. severe**. The symptoms “not eating or drinking”, “conjunctivitis” and “skin lesions” were added in the second wave of the survey (smaller sample size for these symptoms).

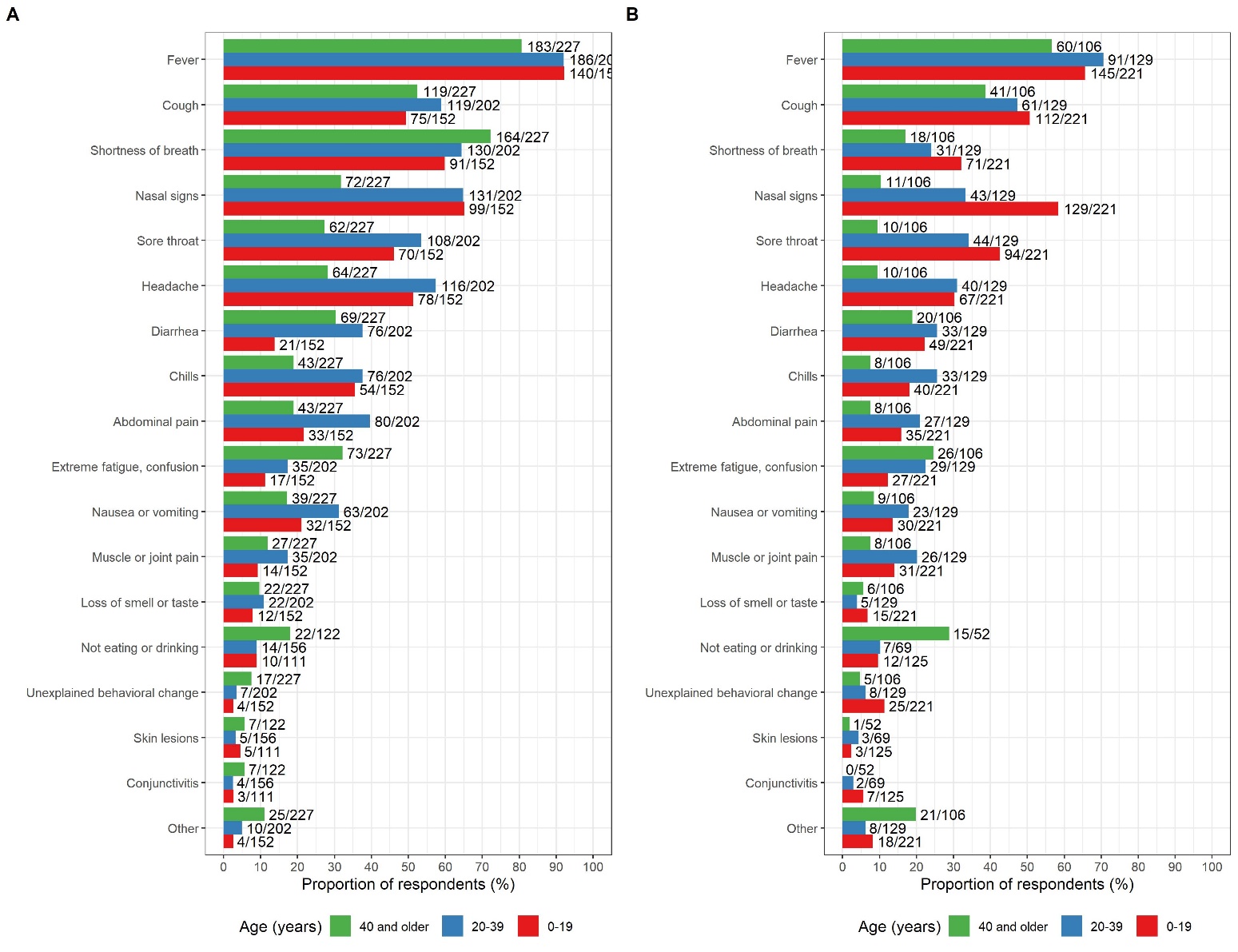

**Figure S5. Signs and symptoms reported among the COVID-19 cases with Down syndrome (T21RS survey) grouped by age and stratified by: A. admitted to hospital, B. not admitted to hospital.** The symptoms “not eating or drinking”, “conjunctivitis” and “skin lesions” were added in the second wave of the survey (smaller sample size for these symptoms).

| **Table S5.** Medical complications grouped by severe outcome of disease (death) (T21RS survey) | | | | |
| --- | --- | --- | --- | --- |
|  | **Overall** | **Did not die*** | **Died** | **p-value**^¶^ |
| n^#^ | 598 | 390 | 111 |  |
| Any medical complications due to COVID-19 (%) | 360 (60.2) | 170 (43.6) | 110 (99.1) | <0.001 |
| Viral COVID-19-related pneumonia (%)^ϕ^ | 204 (36.0) | 119 (31.1) | 68 (68.7) | <0.001 |
| Secondary bacterial pneumonia (%)^ϕ^ | 71 (16.8) | 23 ( 7.8) | 26 (56.5) | <0.001 |
| Acute respiratory distress syndrome (%) | 186 (34.1) | 57 (16.1) | 84 (84.8) | <0.001 |
| Septic shock (%) | 54 (10.5) | 17 ( 4.9) | 18 (22.8) | <0.001 |
| Acute kidney injury (%) | 36 ( 6.9) | 14 ( 4.0) | 13 (15.7) | <0.001 |
| Disseminated intravascular coagulation (%) | 42 ( 8.3) | 9 ( 2.6) | 13 (17.8) | <0.001 |
| Rhabdomyolysis (%) | 10 ( 2.0) | 2 ( 0.6) | 2 ( 2.9) | 0.256 |
| Cardiac failure (%) | 19 ( 3.7) | 3 ( 0.9) | 16 (20.3) | <0.001 |
| Hemorrhage (%) | 25 ( 4.9) | 0 ( 0.0) | 24 (30.4) | <0.001 |
| Multiple organ dysfunction syndrome (%) | 41 ( 8.0) | 0 ( 0.0) | 41 (51.2) | <0.001 |
| Other (%) | 51 (10.1) | 33 ( 9.6) | 17 (23.3) | 0.002 |
| *One of the following clinical situations at last evaluation: 1) Recovered from COVID-19, 2) tested positive but still no symptoms, 3) not currently in hospital but with symptoms or 4) other. Cases without information on the clinical situation at last evaluation or who were still in hospital with symptoms were not included.  ^#^COVID-19 cases with information on medical complications (question was only asked in the clinician survey).  ^¶^Differences between the two groups were tested with Fisher’s exact test  ^ϕ^During the first wave of the survey it was only asked for pneumonia without differentiating between viral and bacterial pneumonia. The question on bacterial pneumonia was added in the second wave of the survey. | | | | |

| **Table S6.** Medical complications stratified by age (T21RS survey) | | | | |
| --- | --- | --- | --- | --- |
|  | **Overall** | **0-19**  **years** | **20-39**  **years** | **40 or**  **older** |
| n^#^ | 598 | 158 | 159 | 281 |
| Any medical complications due to COVID-19 (%) | 360 (60.2) | 64 (40.5) | 103 (64.8) | 193 (68.7) |
| Viral COVID-19-related pneumonia (%)^ϕ^ | 204 (36.0) | 23 (15.6) | 47 (30.9) | 134 (50.2) |
| Secondary bacterial pneumonia (%)^ϕ^ | 71 (16.8) | 13 ( 9.8) | 22 (17.2) | 36 (22.1) |
| Acute respiratory distress syndrome (%) | 186 (34.1) | 31 (21.4) | 53 (37.6) | 102 (39.4) |
| Septic shock (%) | 54 (10.5) | 6 ( 4.3) | 26 (18.6) | 22 ( 9.4) |
| Acute kidney injury (%) | 36 ( 6.9) | 4 ( 2.9) | 8 ( 5.8) | 24 (10.0) |
| Disseminated intravascular coagulation (%) | 42 ( 8.3) | 10 ( 7.2) | 13 ( 9.4) | 19 ( 8.2) |
| Rhabdomyolysis (%) | 10 ( 2.0) | 1 ( 0.7) | 1 ( 0.7) | 8 ( 3.5) |
| Cardiac failure (%) | 19 ( 3.7) | 1 ( 0.7) | 3 ( 2.1) | 15 ( 6.4) |
| Hemorrhage (%) | 25 ( 4.9) | 1 ( 0.7) | 1 ( 0.7) | 23 ( 9.8) |
| Multiple organ dysfunction syndrome (%) | 41 ( 8.0) | 3 ( 2.1) | 4 ( 2.9) | 34 (14.4) |
| Other (%) | 51 (10.1) | 6 ( 4.2) | 9 ( 6.6) | 36 (16.0) |
| ^#^COVID-19 cases with information on medical complications (question was only asked in the clinician survey).  ^ϕ^During the first wave of the survey it was only asked for pneumonia without differentiating between viral and bacterial pneumonia. The question on bacterial pneumonia was added in the second wave of the survey. | | | | |

| **Table S7.** Medical complications among patients with DS stratified by country (T21RS survey). | | | | | | | | | |
| --- | --- | --- | --- | --- | --- | --- | --- | --- | --- |
|  | **Overall** | **India** | **United States** | **Spain** | **Brazil** | **United Kingdom** | **France** | **Italy** | **other** |
| n^#^ | 598 | 216 | 88 | 120 | 36 | 25 | 60 | 21 | 18 |
| Medical complications due to COVID-19 (%) | 360 (60.2) | 158 (73.1) | 40 (45.5) | 81 (67.5) | 8 (22.2) | 10 (40.0) | 25 (41.7) | 18 (85.7) | 8 (44.4) |
| Viral COVID-19-related pneumonia (%)^ϕ^ | 204 (36.0) | 21 (10.8) | 36 (41.4) | 79 (66.4) | 6 (16.7) | 7 (30.4) | 23 (39.0) | 17 (85.0) | 5 (33.3) |
| Secondary bacterial pneumonia (%)^ϕ^ | 71 (16.8) | 55 (26.6) | 11 (14.9) | 0 ( 0.0) | 4 (11.8) | 0 ( 0.0) | 0 ( 0.0) | 1 (25.0) | 0 ( 0.0) |
| Acute respiratory distress syndrome (%) | 186 (34.1) | 106 (49.8) | 14 (17.7) | 26 (24.8) | 7 (20.0) | 2 (10.0) | 11 (19.3) | 16 (80.0) | 4 (28.6) |
| Septic shock (%) | 54 (10.5) | 35 (16.9) | 8 (10.8) | 0 ( 0.0) | 2 ( 5.7) | 0 ( 0.0) | 1 ( 2.0) | 7 (41.2) | 0 ( 0.0) |
| Acute kidney injury (%) | 36 ( 6.9) | 14 ( 6.8) | 8 (10.3) | 7 ( 7.1) | 1 ( 2.9) | 1 ( 5.0) | 3 ( 5.9) | 2 (11.8) | 0 ( 0.0) |
| Disseminated intravascular coagulation (%) | 42 ( 8.3) | 39 (18.9) | 0 ( 0.0) | 0 ( 0.0) | 1 ( 2.9) | 1 ( 5.3) | 0 ( 0.0) | 1 (12.5) | 0 ( 0.0) |
| Rhabdomyolysis (%) | 10 ( 2.0) | 8 ( 3.9) | 0 ( 0.0) | 1 ( 1.1) | 0 ( 0.0) | 0 ( 0.0) | 0 ( 0.0) | 1 (11.1) | 0 ( 0.0) |
| Cardiac failure (%) | 19 ( 3.7) | 6 ( 2.9) | 2 ( 2.6) | 3 ( 3.1) | 0 ( 0.0) | 2 ( 9.5) | 0 ( 0.0) | 6 (37.5) | 0 ( 0.0) |
| Hemorrhage (%) | 25 ( 4.9) | 21 (10.2) | 0 ( 0.0) | 1 ( 1.0) | 0 ( 0.0) | 0 ( 0.0) | 0 ( 0.0) | 3 (18.8) | 0 ( 0.0) |
| Multiple organ dysfunction syndrome (%) | 41 ( 8.0) | 30 (14.4) | 2 ( 2.5) | 0 ( 0.0) | 1 ( 2.9) | 1 ( 5.0) | 0 ( 0.0) | 7 (46.7) | 0 ( 0.0) |
| Other (%) | 51 (10.1) | 0 ( 0.0) | 13 (17.8) | 25 (25.8) | 1 ( 2.9) | 2 ( 9.5) | 2 ( 4.1) | 6 (35.3) | 1 ( 9.1) |
| ^#^COVID-19 cases with information on medical complications (question was only asked in the clinician survey).  ^¶^Differences between the two groups were tested with Fisher’s exact test  ^ϕ^During the first wave of the survey it was only asked for pneumonia without differentiating between viral and bacterial pneumonia. The question on bacterial pneumonia was added in the second wave of the survey. | | | | | | | | | |

| **Table S8.** Study characteristics and comorbidities of the T21RS study sample grouped by severe outcome of disease (death) | | | |
| --- | --- | --- | --- |
|  | **Overall** | **Did not die*** | **Died** |
| n | 1046 | 728 | 131 |
| Age (mean (SD)) | 29.35 (17.92) | 26.68 (17.26) | 50.55 (11.67) |
| Male (%) | 564 (54.0) | 368 (50.6) | 84 (64.6) |
| Family survey (%) | 455 (43.5) | 356 (48.9) | 17 (13.0) |
| Country (%) |  |  |  |
| India | 405 (39.7) | 235 (33.2) | 44 (33.8) |
| Spain | 163 (16.0) | 132 (18.7) | 15 (11.5) |
| United States | 155 (15.2) | 123 (17.4) | 28 (21.5) |
| United Kingdom | 75 ( 7.4) | 66 ( 9.3) | 3 ( 2.3) |
| France | 72 ( 7.1) | 52 ( 7.4) | 10 ( 7.7) |
| Brazil | 67 ( 6.6) | 50 ( 7.1) | 12 ( 9.2) |
| Italy | 35 ( 3.4) | 15 ( 2.1) | 16 (12.3) |
| other | 47 ( 4.6) | 34 ( 4.8) | 2 ( 1.5) |
| Living situation before COVID-19 outbreak (%) |  |  |  |
| Living at home with family | 712 (73.3) | 513 (73.4) | 47 (44.3) |
| Living alone with support | 7 ( 0.7) | 6 ( 0.9) | 0 ( 0.0) |
| Small group home with support | 89 ( 9.2) | 67 ( 9.6) | 14 (13.2) |
| Residential care facility | 157 (16.2) | 108 (15.5) | 44 (41.5) |
| Other | 7 ( 0.7) | 5 ( 0.7) | 1 ( 0.9) |
| Level of intellectual disability (%) |  |  |  |
| Borderline/normal/mild | 169 (18.1) | 137 (20.9) | 7 ( 6.5) |
| Moderate | 580 (62.2) | 412 (63.0) | 54 (50.5) |
| Severe/Profound | 184 (19.7) | 105 (16.1) | 46 (43.0) |
| Admitted to hospital (%) | 581 (56.0) | 306 (42.2) | 117 (90.7) |
| Number of comorbidities (mean (SD)) | 3.81 (2.55) | 3.54 (2.30) | 4.35 (2.86) |
| Obesity (%)^#^ | 342 (38.0) | 224 (35.2) | 43 (39.1) |
| Alzheimer disease/dementia (%)^#^ | 146 (16.2) | 66 (10.4) | 63 (56.2) |
| Thyroid disorder (%)^#^ | 480 (50.8) | 325 (48.4) | 74 (65.5) |
| Seizures/epilepsy (%)^#^ | 233 (25.4) | 132 (20.3) | 47 (42.7) |
| Blood cancer (%) | 17 ( 1.9) | 7 ( 1.1) | 6 ( 5.6) |
| Other cancer (%) | 9 ( 1.0) | 4 ( 0.6) | 4 ( 3.6) |
| Immuno-compromised (%) | 35 ( 3.9) | 20 ( 3.1) | 9 ( 8.3) |
| Obstructive sleep apnea (%)^#^ | 311 (34.6) | 220 (34.4) | 21 (20.2) |
| Hypertension (%) | 106 (11.7) | 54 ( 8.5) | 19 (17.4) |
| Diabetes (%)^#^ | 195 (21.5) | 107 (16.7) | 26 (24.3) |
| Cerebrovascular disease (%) | 22 ( 2.5) | 15 ( 2.4) | 5 ( 4.9) |
| Coronary heart disease (%) | 67 ( 7.4) | 42 ( 6.6) | 10 ( 9.2) |
| Chronic renal disease (%) | 79 ( 8.8) | 46 ( 7.3) | 12 (11.1) |
| Chronic liver disease (%) | 153 (17.2) | 93 (14.8) | 13 (12.0) |
| Chronic lung disease (%)^#^ | 265 (29.3) | 174 (27.3) | 30 (27.5) |
| Celiac disease (%) | 60 ( 6.7) | 48 ( 7.6) | 3 ( 2.8) |
| Gastroesophageal reflux (GERD) (%)^#^ | 223 (24.8) | 152 (23.9) | 25 (22.7) |
| Irritable bowel syndrome (IBS) (%) | 118 (13.2) | 74 (11.7) | 9 ( 8.2) |
| Hepatitis B infection (%) | 23 ( 2.6) | 14 ( 2.3) | 6 ( 5.6) |
| Congenital heart defect (%)^#^ | 392 (40.2) | 276 (39.7) | 29 (25.4) |
| Behavioral or psychiatric condition (%)^#^ | 465 (48.2) | 296 (42.9) | 63 (58.3) |
| Other pre-existing conditions (%) | 249 (26.7) | 188 (28.2) | 53 (49.1) |
| *One of the following clinical situations at last evaluation: 1) Recovered from COVID-19, 2) tested positive but still no symptoms, 3) not currently in hospital but with symptoms or 4) other. Cases without information on the clinical situation at last evaluation or who were still in hospital with symptoms were not included. ^#^Comorbidities that were present in at least 15% of our analysis sample were included in the subsequent association analyses. | | | |

| **Table S9.** Age distribution of the proportion of deaths among individuals with DS (T21RS and ISARIC4C surveys), who were hospitalized with COVID-19, in comparison to hospitalized cases of COVID-19 from the general population (data from the UK ISARIC4C survey, NYC^1^ and Spain^2^). | | | | |
| --- | --- | --- | --- | --- |
| **A. Individuals with Down syndrome** | | | | |
| **Population** | **Age group** | **n in age group** | **n of deaths** | **% died** |
| T21RS survey | 0-19 | 100 | 4 | 4.00 |
|  | 20-29 | 61 | 1 | 1.64 |
|  | 30-39 | 63 | 8 | 12.70 |
|  | 40-49 | 70 | 26 | 37.14 |
|  | 50-59 | 97 | 56 | 57.73 |
|  | 60-69 | 29 | 21 | 72.41 |
|  | 70-79 | 2 | 0 | 0.00 |
|  | 80 and older | 0 | 0 | 0.00 |
| ISARIC4C survey | 0-19 | 9 | 0 | 0.00 |
|  | 20-29 | 7 | 2 | 28.57 |
|  | 30-39 | 10 | 2 | 20.00 |
|  | 40-49 | 19 | 8 | 42.11 |
|  | 50-59 | 34 | 14 | 41.18 |
|  | 60-69 | 24 | 14 | 58.33 |
|  | 70-79 | 5 | 3 | 60.00 |
|  | 80 and older | 0 | 0 | 0.00 |
| **B. General population** | | | | |
| **Population** | **Age group** | **n in age group** | **n of deaths** | **% died** |
| UK^1^ | 0-19 | 909 | 12 | 1.32 |
|  | 20-29 | 1073 | 22 | 2.05 |
|  | 30-39 | 2113 | 92 | 4.35 |
|  | 40-49 | 3931 | 304 | 7.73 |
|  | 50-59 | 7163 | 1056 | 14.74 |
|  | 60-69 | 8754 | 2340 | 26.73 |
|  | 70-79 | 13095 | 5059 | 38.63 |
|  | 80 and older | 21388 | 10255 | 47.95 |
| Spain | 0-19 | 0 | 0 | NA |
|  | 20-29 | 159 | 1 | 0.63 |
|  | 30-39 | 219 | 0 | 0.00 |
|  | 40-49 | 287 | 4 | 1.39 |
|  | 50-59 | 365 | 14 | 3.84 |
|  | 60-69 | 272 | 36 | 13.24 |
|  | 70-79 | 358 | 122 | 34.08 |
|  | 80 and older | 511 | 283 | 55.38 |
| New York City | 0-19 | 34 | 0 | 0.00 |
|  | 20-29 | 97 | 4 | 4.12 |
|  | 30-39 | 211 | 8 | 3.79 |
|  | 40-49 | 352 | 22 | 6.25 |
|  | 50-59 | 515 | 53 | 10.29 |
|  | 60-69 | 533 | 84 | 15.76 |
|  | 70-79 | 451 | 145 | 32.15 |
|  | 80 and older | 441 | 237 | 53.74 |
| ^1^individuals without Down syndrome from the ISARIC4C survey | | | | |

| **Table S10. Mortality rates among patients hospitalized with COVID-19 stratified by age.** Hospitalized individuals with Down syndrome from the UK ISARIC4C and the T21RS surveys are compared to matched individuals without Down syndrome (controls) from the ISARIC4C survey. The 100 individuals with Down syndrome reported through the UK ISARIC4C survey were matched to 400 individuals without Down syndrome (controls) from the same survey (matching 1:4) as well as to 100 individuals with Down syndrome from the T21RS survey (matching 1:1). As samples were matched based on age, gender and ethnicity, mortality rates are corrected for these characteristics (see supplementary tables S2 and S3 for more information on the matched samples). We had information on the outcome of disease in 88/100 individuals from the matched T21RS dataset. | | | | | | |
| --- | --- | --- | --- | --- | --- | --- |
| **A. All matched individuals** | | | | | | |
|  | **ISARIC4C Controls** | | **ISARIC4C individuals**  **with Down syndrome** | | **T21RS matched individuals**  **with Down syndrome** | |
|  | **Yes** | **No** | **Yes** | **No** | **Yes** | **No** |
| **Died** | 55 (13.7%) | 345 (86.3%) | 40 (40.0%) | 60 (60.0%) | 42 (47.7%) | 46 (52.3%) |
| **B.** **Younger than 40 years of age** | | | | | | |
|  | **Yes** | **No** | **Yes** | **No** | **Yes** | **No** |
| **Died** | 3 (3.0%) | 97 (97.0%) | 3 (12.0%) | 22 (88.0%) | 2 (12.5%) | 14 (87.5%) |
| **C.** **40 years of age or older** | | | | | | |
|  | **Yes** | **No** | **Yes** | **No** | **Yes** | **No** |
| **Died** | 52 (17.3%) | 248 (82.7%) | 37 (49.3%) | 38 (50.7%) | 40 (55.6%) | 32 (44.4%) |

| **Table S11. Association between Down syndrome and mortality after hospitalization with COVID-19 stratified by age.** Hospitalized individuals with Down syndrome from the UK ISARIC4C and the T21RS surveys are compared to matched individuals without Down syndrome (controls) from the ISARIC4C survey. The 100 individuals with Down syndrome reported through the UK ISARIC4C survey were matched to 400 individuals without Down syndrome (controls) from the same survey (matching 1:4) as well as to 100 individuals with Down syndrome from the T21RS survey (matching 1:1). As samples were matched based on age, gender and ethnicity, mortality rates are corrected for these characteristics (see supplementary tables S2 and S3 for more information on the matched samples). We had information on the outcome of disease in 88/100 individuals from the matched T21RS dataset. | | | |
| --- | --- | --- | --- |
| **A. All individuals** | | | |
|  | **RR** | **95% Confidence intervals** | ***p-value*** |
| ***Adjusted for age, gender, ethnicity*** | | | |
| Matched T21RS vs. ISARIC4C controls | 3.47 | 2.58 – 4.39 | **<0.0001** |
| Matched samples within ISARIC4C | 2.91 | 2.11 – 3.79 | **< 0.0001** |
| All samples within ISARIC4C | 2.17 | 1.88 – 2.42 | **< 0.0001** |
| ***Additionally adjusted for known risk factors for***  ***mortality^1^*** | | | |
| Matched samples within ISARIC4C | 2.49 | 1.51 – 3.69 | **0.0006** |
| All samples within ISARIC4C | 2.04 | 1.65 – 2.36 | **< 0.0001** |
| **B. Younger than 40 years of age** | | | |
|  | **RR** | **95% Confidence intervals** | ***p-value*** |
| ***Adjusted for age, gender, ethnicity*** |  |  |  |
| Matched T21RS vs. ISARIC4C controls | 4.17 | 0.58 – 16.13 | 0.11 |
| Matched samples within ISARIC4C | 4.00 | 0.78 – 14.62 | 0.0809 |
| All samples within ISARIC4C | 4.01 | 1.02 – 9.74 | **0.0194** |
| ***Additionally adjusted for known risk factors for mortality^1^*** |  |  |  |
| Matched samples within ISARIC4C | 2.42 | 0.12 – 12.88 | 0.4370 |
| All samples within ISARIC4C^2^ | 1.42 | 0.08 – 6.30 | 0.7320 |
| **C. 40 years of age or older** | | | |
|  | **RR** | **95% Confidence intervals** | ***p-value*** |
| ***Adjusted for age, gender, ethnicity*** |  |  |  |
| Matched T21RS vs. ISARIC4C controls | 3.21 | 2.42 – 3.96 | **<0.0001** |
| Matched samples within ISARIC4C | 2.85 | 2.09 – 3.62 | **< 0.0001** |
| All samples within ISARIC4C | 2.44 | 2.01 – 2.80 | **< 0.0001** |
| ***Additionally adjusted for known risk factors for mortality^1^*** |  |  |  |
| Matched samples within ISARIC4C | 2.73 | 1.71–3.84 | **0.0001** |
| All samples within ISARIC4C | 2.26 | 1.74 – 2.73 | **< 0.0001** |
| ^1^Chronic cardiac disease, chronic pulmonary disease, chronic kidney disease, liver disease, obesity, chronic neurological disorder, dementia, malignant neoplasm (from reference ^3^)  ^2^Dementia and malignant neoplasm were removed from the model as these were not present in this age group | | | |

| **Table S12. Association between age (younger than 40 years of age versus 40 years or older (reference)) and mortality after hospitalization with COVID-19.** | | | | |
| --- | --- | --- | --- | --- |
| **A. T21RS (hospitalized patients)** | | | | |
|  | **N** | **Reduced risk for mortality^1^ (%)** | **95% Confidence intervals** | ***p-value*** |
| ***Adjusted for gender, data source (family vs. clinician survey), country*** | | | | |
| Younger than 40 years of age versus 40 years or older (reference) | 417 | 92.50 | 85.12 – 96.57 | **<0.0001** |
| ***Additionally adjusted for number of DS related comorbidities^2^ and number of general risk factors^3^*** | | | | |
| Younger than 40 years of age versus 40 years or older (reference) | 417 | 91.68 | 83.54 – 96.20 | **<0.0001** |
| **B. ISARIC4C (hospitalized patients with Down syndrome)** | | | | |
|  | **N** | **Reduced risk for mortality^1^ (%)** | **95% Confidence intervals** | ***p-value*** |
| ***Adjusted for gender and ethnicity*** | | | | |
| Younger than 40 years of age versus 40 years or older (reference) | 100 | 72.68 | 29.07 – 93.69 | **0.00867** |
| ***Additionally adjusted for number of DS related comorbidities^4^ and number of general risk factors^5^*** | | | | |
| Younger than 40 years of age versus 40 years or older (reference) | 100 | 67.61 | 18.12 – 92.50 | **0.0215** |
| ^1^Calculated as (1 - RR)*100  ^2^Thyroid disorder, Seizures/epilepsy, Obstructive sleep apnea, gastroesophageal reflux (GERD), congenital heart defect, behavioral and psychiatric condition  ^3^Obesity, dementia, blood cancer, other cancer, immuno-compromised, hypertension, diabetes, coronary heart disease, kidney, liver, lung diseases  ^4^Sleep apnea, reflux and dysphagia, behavior and psychiatric conditions, thyroid disorder, rheumatologic disorder  ^5^Chronic cardiac disease, dementia, chronic pulmonary disease (not asthma), asthma, chronic kidney disease, mild, moderate or severe liver disease, chronic neurological disorder, chronic hematologic disease, obesity, diabetes, smoking | | | | |

| **Table S13.** Risk factors associated with adverse outcomes of COVID-19 in individuals with Down syndrome. Associations with hospitalization and mortality in symptomatic COVID-19 patients with Down syndrome from the T21RS survey estimated in adjusted logistic regression models (odds ratios (OR) and 95%-confidence intervals (95%-CI)). | | | | | | |
| --- | --- | --- | --- | --- | --- | --- |
|  | **Hospitalization** | | | **Mortality** | | |
| **Risk factors** | **OR (95%-CI)** | **p-value** | **N** | **OR (95%-CI)** | **p-value** | **N** |
| Age | 1.75 (1.47; 2.07) | 5.24E-08 | 975 | 2.41 (2.02; 2.89) | 7.08E-22 | 802 |
| Male | 1.51 (1.16; 1.97) | 0.002 | 973 | 1.75 (1.16; 2.63) | 0.007 | 800 |
| Living in residential care facility | 0.85 (0.50; 1.45) | 0.556 | 878 | 1.18 (0.61; 2.29) | 0.617 | 725 |
| Level of IDD: borderline/normal/mild (ref) | 1.00 (1.00; 1.00) | NA | 852 | 1.00 (1.00; 1.00) | NA | 694 |
| Level of IDD: moderate | 1.21 (0.78; 1.89) | 0.400 | 852 | 0.81 (0.30; 2.17) | 0.676 | 694 |
| Level of IDD: severe/profound | 1.19 (0.67; 2.09) | 0.552 | 852 | 1.33 (0.47; 3.77) | 0.591 | 694 |
| Number comorbidities | 1.12 (0.90; 1.41) | 0.319 | 950 | 1.26 (0.89; 1.77) | 0.189 | 780 |
| Obesity | 2.03 (1.44; 2.87) | 0.000 | 829 | 1.33 (0.75; 2.35) | 0.323 | 684 |
| Alzheimer disease/dementia | 0.77 (0.44; 1.36) | 0.372 | 833 | 2.13 (1.10; 4.12) | 0.025 | 688 |
| Thyroid disorder | 1.10 (0.80; 1.53) | 0.551 | 863 | 1.23 (0.69; 2.17) | 0.482 | 714 |
| Seizures/epilepsy | 1.21 (0.81; 1.79) | 0.351 | 846 | 1.61 (0.89; 2.93) | 0.117 | 699 |
| Obstructive sleep apnea | 1.17 (0.84; 1.65) | 0.351 | 829 | 0.68 (0.37; 1.26) | 0.224 | 684 |
| Gastroesophageal reflux | 0.91 (0.62; 1.34) | 0.638 | 830 | 1.28 (0.63; 2.62) | 0.494 | 686 |
| Congenital heart defect | 1.46 (1.05; 2.03) | 0.026 | 886 | 0.89 (0.47; 1.66) | 0.704 | 735 |
| Behavioral and psychiatric condition | 1.15 (0.81; 1.62) | 0.446 | 876 | 0.85 (0.48; 1.49) | 0.563 | 723 |
| Chronic lung disease | 0.89 (0.60; 1.31) | 0.546 | 836 | 0.80 (0.38; 1.70) | 0.562 | 688 |
| Diabetes | 1.93 (1.20; 3.12) | 0.007 | 837 | 0.54 (0.24; 1.21) | 0.136 | 689 |
| Associations with age and gender were adjusted for the data source (caregiver versus clinician survey) and associations with living situation, level of IDD and comorbidities were adjusted for age, gender, data source and country of residence. Abbreviations: IDD, intellectual and developmental disabilities; ref, reference category | | | | | | |
