## Supplementary material for "An international survey on the impact of COVID-19 in individuals with Down syndrome": Author group (T21RS COVID-19 Initiative)

| **First Initials** | **Surname** | **Affiliation** |
| --- | --- | --- |
| TdJ | Bermejo | Instituto Hispalense de Psiquiatría, Spain |
| P | Borrell | DOWN ESPANA, Barcelona, Spain |
| L | Cretu | Institut Jérôme Lejeune, Paris, France |
| R | de la Torre | Hospital del Mar Medical Research Institute, Spain |
| J | Florez | Fundacion Iberoamericana Down 21, Spain |
| J | Fortea | Sant Pau Memory Unit, Department of Neurology, Hospital de la Santa Creu i Sant Pau, Biomedical Research Institute Sant Pau, Universitat Autònoma de Barcelona, Barcelona, Spain; Barcelona Down Medical Center, Fundació Catalana de Síndrome de Down, Barcelona, Spain |
| P | Ghosh | Department of Zoology. Bijoy Krishna Girls' College. Howrah, West Bengal, India |
| D | González-Lamuño | Hospital Valdecilla, Santander, Cantabria, Spain |
| A | Hiance-Delahaye | Institut Jérôme Lejeune, Paris, France |
| C | Laffon | Institut Jérôme Lejeune, Paris, France |
| J | Levin | Department of Neurology, German Center of Neurodegenerative Diseases; Ludwig-Maximilians-Universität (LMU); Munich Cluster for Systems Neurology (SyNergy), Munich, Germany |
| A | Matia | DOWN ESPANA, Barcelona, Spain |
| C | Mircher | Institut Jérôme Lejeune, Paris, France |
| F | Moldenhauer | Hospital La Princesa, Madrid, Spain |
| M | Mulqueen | Advantage Care Health Centers, Brookville, NY, USA |
| NJ | Nowalk | Institut Jérôme Lejeune, Paris, France |
| E | Prioux | Research Programme on Biomedical Informatics, Hospital del Mar and DCEXS Universitat Pompeu Fabra, Barcelona, Spain |
| F | Sanz | Institut Jérôme Lejeune, Paris, France |
| J | Toulas | Children’s Hospital of Pittsburgh, Pittsburgh, PA, USA |
