## Supplementary material for "An international survey on the impact of COVID-19 in individuals with Down syndrome": Caregiver/family survey

Emory University

Library Information Technology Services

**Down syndrome and COVID-19**

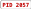

$ Codebook

**Data Dictionary Codebook**

**Caregiver/family Survey—English version**	10/25/2020 8:27am

% Collapse all instruments

| 1 | Self-reporting. I am the person with Down syndrome |
| --- | --- |
| 2 | Self-reporting or family member |
| 3 | Unrelated caregiver |
| 4 | Other |

| **#** | **Variable / Field Name** | **Field Label**  ***Field Note*** | **Field Attributes (Field Type, Validation, Choices, Calculations, etc.)** | | | | |
| --- | --- | --- | --- | --- | --- | --- | --- |
| Instrument: **Landing Page** (landing_page) ' Enabled as survey & Expand | | | | | | | |
| Instrument: **Family Eng** (family_eng) ' Enabled as survey % Collapse | | | | | | | |
| 11 | relation | Section Header: *Please describe who you are and then answer general questions about the person with Down syndrome.*  What is your relationship to the person with Down syndrome? | radio  Field Annotation: @HIDECHOICE="1" | | | | |
| 12 | other_relation  Show the ﬁeld ONLY if: [relation] = '4' | Please specify your relationship. | text | | | | |
| 13 | age | How old (in years) is the person with Down syndrome? (Please enter "0" if the person is a newborn baby.) | text (number, Min: 0, Max: 100), Required | | | | |
| 14 | gender | How do they identify themselves? | radio, Required | | | | |
|  |  |  |  | 1 | Female | | |
|  |  |  |  | 2 | Male | | |
|  |  |  |  | 3 | Other | | |
|  |  |  |  | 4 | Choose not to answer | | |
| 15 | race | What is their race? | text  Field Annotation: @HIDDEN | | | | |
| 16 | who_group | What is their ethnic group (as deﬁned by World Health  Organization)? Please check all that apply: | checkbox | | | | |
|  |  |  |  | 1 | | who_group 1 | Aboriginal First Nations |
|  |  |  |  | 2 | | who_group 2 | Arab |
|  |  |  |  | 3 | | who_group 3 | Black |
|  |  |  |  | 4 | | who_group 4 | East Asian (e.g., China, Korea, Japan) |
|  |  |  |  | 5 | | who_group 5 | Latin American (e.g., Mexico, Brazil, Peru) |
|  |  |  |  | 6 | | who_group 6 | South Asian (e.g., Afghanistan, India, Pakistan) |
|  |  |  |  | 7 | | who_group 7 | West Asian (e.g., Saudi  Arabia, Syria, Turkey) |
|  |  |  |  | 8 | | who_group 8 | White |
|  |  |  |  | 9 | | who_group 9 | Other |
|  |  |  |  | 10 | | who_group 10 | Choose not to answer |
| 17 | ethnicity | How do they describe their ethnicity? | text | | | | |

| 1 | Borderline/normal |
| --- | --- |
| 2 | Mild |
| 3 | Moderate |
| 4 | Severe/Profound |
| 5 | Don't know |
| 6 | Choose not to answer |

| 18 | trisomy_type | Type of trisomy 21 | radio | | |
| --- | --- | --- | --- | --- | --- |
|  |  |  |  | 1 | Full/standard |
|  |  |  |  | 2 | Mosaic |
|  |  |  |  | 3 | Translocation |
|  |  |  |  | 4 | Partial trisomy |
|  |  |  |  | 5 | Don't know |
| 19 | id_level | Level of intellectual disability | radio  Field Annotation: @HIDECHOICE="1" | | |

20	country	Section Header:

| 1 | Afghanistan |
| --- | --- |
| 2 | Albania |
| 3 | Algeria |
| 4 | Andorra |
| 5 | Angola |
| 6 | Antigua & Deps |
| 7 | Argentina |
| 8 | Armenia |
| 9 | Australia |
| 10 | Austria |
| 11 | Azerbaijan |
| 12 | Bahamas |
| 13 | Bahrain |
| 14 | Bangladesh |
| 15 | Barbados |
| 16 | Belarus |
| 17 | Belgium |
| 18 | Belize |
| 19 | Benin |
| 20 | Bhutan |
| 21 | Bolivia |
| 22 | Bosnia Herzegovina |
| 23 | Botswana |
| 24 | Brazil |
| 25 | Brunei |
| 26 | Bulgaria |
| 27 | Burkina |
| 28 | Burundi |
| 29 | Cambodia |
| 30 | Cameroon |
| 31 | Canada |
| 32 | Cape Verde |
| 33 | Central African Rep |
| 34 | Chad |
| 35 | Chile |
| 36 | China |
| 37 | Colombia |
| 38 | Comoros |
| 39 | Congo |
| 40 | Congo {Democratic Rep} |
| 41 | Costa Rica |
| 42 | Croatia |
| 43 | Cuba |
| 44 | Cyprus |
| 45 | Czech Republic |
| 46 | Denmark |
| 47 | Djibouti |
| 48 | Dominica |

In what country do they live?

dropdown

|  |  |  |  | 49 | Dominican Republic |
| --- | --- | --- | --- | --- | --- |
|  |  |  |  | 50 | East Timor |
|  |  |  |  | 51 | Ecuador |
|  |  |  |  | 52 | Egypt |
|  |  |  |  | 53 | El Salvador |
|  |  |  |  | 54 | Equatorial Guinea |
|  |  |  |  | 55 | Eritrea |
|  |  |  |  | 56 | Estonia |
|  |  |  |  | 57 | Ethiopia |
|  |  |  |  | 58 | Fiji |
|  |  |  |  | 59 | Finland |
|  |  |  |  | 60 | France |
|  |  |  |  | 61 | Gabon |
|  |  |  |  | 62 | Gambia |
|  |  |  |  | 63 | Georgia |
|  |  |  |  | 64 | Germany |
|  |  |  |  | 65 | Ghana |
|  |  |  |  | 66 | Greece |
|  |  |  |  | 67 | Grenada |
|  |  |  |  | 68 | Guatemala |
|  |  |  |  | 69 | Guinea |
|  |  |  |  | 70 | Guinea-Bissau |
|  |  |  |  | 71 | Guyana |
|  |  |  |  | 72 | Haiti |
|  |  |  |  | 73 | Honduras |
|  |  |  |  | 74 | Hungary |
|  |  |  |  | 75 | Iceland |
|  |  |  |  | 76 | India |
|  |  |  |  | 77 | Indonesia |
|  |  |  |  | 78 | Iran |
|  |  |  |  | 79 | Iraq |
|  |  |  |  | 80 | Ireland {Republic} |
|  |  |  |  | 81 | Israel |
|  |  |  |  | 82 | Italy |
|  |  |  |  | 83 | Ivory Coast |
|  |  |  |  | 84 | Jamaica |
|  |  |  |  | 85 | Japan |
|  |  |  |  | 86 | Jordan |
|  |  |  |  | 87 | Kazakhstan |
|  |  |  |  | 88 | Kenya |
|  |  |  |  | 89 | Kiribati |
|  |  |  |  | 90 | Korea North |
|  |  |  |  | 91 | Korea South |
|  |  |  |  | 92 | Kosovo |
|  |  |  |  | 93 | Kuwait |
|  |  |  |  | 94 | Kyrgyzstan |
|  |  |  |  | 95 | Laos |
|  |  |  |  | 96 | Latvia |

|  |  |  |  | 97 | Lebanon |
| --- | --- | --- | --- | --- | --- |
|  |  |  |  | 98 | Lesotho |
|  |  |  |  | 99 | Liberia |
|  |  |  |  | 100 | Libya |
|  |  |  |  | 101 | Liechtenstein |
|  |  |  |  | 102 | Lithuania |
|  |  |  |  | 103 | Luxembourg |
|  |  |  |  | 104 | Macedonia |
|  |  |  |  | 105 | Madagascar |
|  |  |  |  | 106 | Malawi |
|  |  |  |  | 107 | Malaysia |
|  |  |  |  | 108 | Maldives |
|  |  |  |  | 109 | Mali |
|  |  |  |  | 110 | Malta |
|  |  |  |  | 111 | Marshall Islands |
|  |  |  |  | 112 | Mauritania |
|  |  |  |  | 113 | Mauritius |
|  |  |  |  | 114 | Mexico |
|  |  |  |  | 115 | Micronesia |
|  |  |  |  | 116 | Moldova |
|  |  |  |  | 117 | Monaco |
|  |  |  |  | 118 | Mongolia |
|  |  |  |  | 119 | Montenegro |
|  |  |  |  | 120 | Morocco |
|  |  |  |  | 121 | Mozambique |
|  |  |  |  | 195 | Myanmar (Burma) |
|  |  |  |  | 122 | Namibia |
|  |  |  |  | 123 | Nauru |
|  |  |  |  | 124 | Nepal |
|  |  |  |  | 125 | Netherlands |
|  |  |  |  | 126 | New Zealand |
|  |  |  |  | 127 | Nicaragua |
|  |  |  |  | 128 | Niger |
|  |  |  |  | 129 | Nigeria |
|  |  |  |  | 130 | Norway |
|  |  |  |  | 131 | Oman |
|  |  |  |  | 132 | Pakistan |
|  |  |  |  | 133 | Palau |
|  |  |  |  | 134 | Panama |
|  |  |  |  | 135 | Papua New Guinea |
|  |  |  |  | 136 | Paraguay |
|  |  |  |  | 137 | Peru |
|  |  |  |  | 138 | Philippines |
|  |  |  |  | 139 | Poland |
|  |  |  |  | 140 | Portugal |
|  |  |  |  | 141 | Qatar |
|  |  |  |  | 142 | Romania |
|  |  |  |  | 143 | Russian Federation |

|  |  |  |  | 144 | Rwanda |
| --- | --- | --- | --- | --- | --- |
|  |  |  |  | 145 | St Kitts & Nevis |
|  |  |  |  | 146 | St Lucia |
|  |  |  |  | 147 | Saint Vincent & the Grenadines |
|  |  |  |  | 148 | Samoa |
|  |  |  |  | 149 | San Marino |
|  |  |  |  | 150 | Sao Tome & Principe |
|  |  |  |  | 151 | Saudi Arabia |
|  |  |  |  | 152 | Senegal |
|  |  |  |  | 153 | Serbia |
|  |  |  |  | 154 | Seychelles |
|  |  |  |  | 155 | Sierra Leone |
|  |  |  |  | 156 | Singapore |
|  |  |  |  | 157 | Slovakia |
|  |  |  |  | 158 | Slovenia |
|  |  |  |  | 159 | Solomon Islands |
|  |  |  |  | 160 | Somalia |
|  |  |  |  | 161 | South Africa |
|  |  |  |  | 162 | Spain |
|  |  |  |  | 163 | Sri Lanka |
|  |  |  |  | 164 | Sudan |
|  |  |  |  | 165 | Suriname |
|  |  |  |  | 166 | Swaziland |
|  |  |  |  | 167 | Sweden |
|  |  |  |  | 168 | Switzerland |
|  |  |  |  | 169 | Syria |
|  |  |  |  | 170 | Taiwan |
|  |  |  |  | 171 | Tajikistan |
|  |  |  |  | 172 | Tanzania |
|  |  |  |  | 173 | Thailand |
|  |  |  |  | 174 | Togo |
|  |  |  |  | 175 | Tonga |
|  |  |  |  | 176 | Trinidad & Tobago |
|  |  |  |  | 177 | Tunisia |
|  |  |  |  | 178 | Turkey |
|  |  |  |  | 179 | Turkmenistan |
|  |  |  |  | 180 | Tuvalu |
|  |  |  |  | 181 | Uganda |
|  |  |  |  | 182 | Ukraine |
|  |  |  |  | 183 | United Arab Emirates |
|  |  |  |  | 184 | United Kingdom |
|  |  |  |  | 185 | United States |
|  |  |  |  | 186 | Uruguay |
|  |  |  |  | 187 | Uzbekistan |
|  |  |  |  | 188 | Vanuatu |
|  |  |  |  | 189 | Vatican City |
|  |  |  |  | 190 | Venezuela |
|  |  |  |  | 191 | Vietnam |

192	Yemen

193	Zambia

194	Zimbabwe

21	region	What region of that country (e.g., state, province, district, county)?

text

22	residence	What best describes their living situation before the COVID-19 outbreak?

radio

1	Living at home with family

7	Living alone with no support (autonomously)

2	Living alone with support

3	Small group home with support

4	Residential care facility

5	Other

6	Don't know

23	residence_other

Show the ﬁeld ONLY if: [residence] = '5'

Please specify other setting.	text

24	change_residence	Did they change their living situation because of the COVID-19 outbreak?

radio

1	Yes

2	No

3	Don't know

25	change

Show the ﬁeld ONLY if: [change_residence] = '1'

Please describe their current living situation.	radio

1	Living at home with family

7	Living alone with no support (autonomously)

2	Living alone with support

3	Small group home with support

4	Residential care facility

5	Other

6	Don't know / Not applicable

26	change_oth

Show the ﬁeld ONLY if: [change] = '5'

Please specify other setting.	text

27	contacts_yn	Did anyone test positive or have signs or symptoms of

COVID-19 where the person with Down syndrome lived during the outbreak?

radio

1	Yes

2	No

3	Don't know / Not applicable

28	chd_yn	Section Header: *This section asks about medical conditions that existed before the COVID-19 outbreak (referred to as "pre-existing conditions").*

Did the person with Down syndrome have a congenital heart defect?

radio

1	Yes

2	No

3	Don't know

29	chd_surgery

Show the ﬁeld ONLY if: [chd_yn] = '1'

Did the heart defect require surgery?	radio

1	Yes surgery was done and fully correct the defect

2	Yes surgery was done but did not fully correct the defect

3	No surgery was required

4	Don't know

30	psychiatric_yn	Does the person with Down syndrome have a current diagnosis of any behavioral and psychiatric condition (for example,

| 1 | Yes |
| --- | --- |
| 2 | No |
| 3 | Don't know |

autism spectrum disorder)?

radio

31	psychiatric_type

Show the ﬁeld ONLY if: [psychiatric_yn] = '1'

Please check all that apply.	checkbox

1	psychiatric_type 1	Autism spectrum disorder

2	psychiatric_type 2	Depression

3	psychiatric_type 3	ADHD/ADD

4	psychiatric_type 4	Anxiety

5	psychiatric_type 5	Obsessive-compulsive disorder (OCD)

6	psychiatric_type 6	Behavior problems

7	psychiatric_type 7	Psychosis

8	psychiatric_type 8	Other (for example, regression disorder/disintegrative disorder, ...)

32	psych_other

Please specify.	text

Show the ﬁeld ONLY if: [psychiatric_type(8)] = '1'

33	alz	Section Header: *Please check the following pre-existing conditions diagnosed by a doctor if experienced by the person with Down syndrome.*

Alzheimer disease/dementia

radio (Matrix)

1	Yes--currently being treated

2	Yes--not being treated

3	In the past--no current treatment

4	Never diagnosed

5	Don't know

34	thyroid	Thyroid disorder	radio (Matrix)

1	Yes--currently being treated

2	Yes--not being treated

3	In the past--no current treatment

4	Never diagnosed

5	Don't know

35	seizure	Seizures/epilepsy	radio (Matrix)

1	Yes--currently being treated

2	Yes--not being treated

3	In the past--no current treatment

4	Never diagnosed

5	Don't know

36	blood_cancer	Blood cancer (e.g., leukemia,lymphoma)	radio (Matrix)

1	Yes--currently being treated

2	Yes--not being treated

3	In the past--no current treatment

4	Never diagnosed

5	Don't know

37	other_cancer	Other cancer	radio (Matrix)

| 1 | Yes--currently being treated |
| --- | --- |
| 2 | Yes--not being treated |
| 3 | In the past--no current treatment |
| 4 | Never diagnosed |
| 5 | Don't know |

38	immumo	Immuno-compromised (e.g,, on cancer treatment)	radio (Matrix)

1	Yes--currently being treated

2	Yes--not being treated

3	In the past--no current treatment

4	Never diagnosed

5	Don't know

39	osa	Obstructive sleep apnea	radio (Matrix)

1	Yes--currently being treated

2	Yes--not being treated

3	In the past--no current treatment

4	Never diagnosed

5	Don't know

40	obesity	Obesity	radio (Matrix)

1	Yes--currently being treated

2	Yes--not being treated

3	In the past--no current treatment

4	Never diagnosed

5	Don't know

41	hyperten	Hypertension	radio (Matrix)

1	Yes--currently being treated

2	Yes--not being treated

3	In the past--no current treatment

4	Never diagnosed

5	Don't know

42	diabetes	Diabetes	radio (Matrix)

1	Yes--currently being treated

2	Yes--not being treated

3	In the past--no current treatment

4	Never diagnosed

5	Don't know

43	cvd	Cerebrovascular disease	radio (Matrix)

1	Yes--currently being treated

2	Yes--not being treated

3	In the past--no current treatment

4	Never diagnosed

5	Don't know

44	cad	Coronary heart disease	radio (Matrix)

1	Yes--currently being treated

2	Yes--not being treated

3	In the past--no current treatment

4	Never diagnosed

5	Don't know

45	renal	Chronic renal disease	radio (Matrix)

| 1 | Yes--currently being treated |
| --- | --- |
| 2 | Yes--not being treated |
| 3 | In the past--no current treatment |
| 4 | Never diagnosed |
| 5 | Don't know |

46	liver	Chronic liver disease	radio (Matrix)

1	Yes--currently being treated

2	Yes--not being treated

3	In the past--no current treatment

4	Never diagnosed

5	Don't know

47	lung	Chronic lung disease (e.g., asthma, emphysema, COPD)	radio (Matrix)

1	Yes--currently being treated

2	Yes--not being treated

3	In the past--no current treatment

4	Never diagnosed

5	Don't know

48	celiac	Celiac disease	radio (Matrix)

1	Yes--currently being treated

2	Yes--not being treated

3	In the past--no current treatment

4	Never diagnosed

5	Don't know

49	gerd	Gastroesophageal reﬂux (GERD)	radio (Matrix)

1	Yes--currently being treated

2	Yes--not being treated

3	In the past--no current treatment

4	Never diagnosed

5	Don't know

50	ibs	Irritable bowel syndrome (IBS)	radio (Matrix)

1	Yes--currently being treated

2	Yes--not being treated

3	In the past--no current treatment

4	Never diagnosed

5	Don't know

51	hep_b	Heaptitis B infection	radio (Matrix)

1	Yes--currently being treated

2	Yes--not being treated

3	In the past--no current treatment

4	Never diagnosed

5	Don't know

52	onset_seiz

Show the ﬁeld ONLY if: [seizure] = '1' or [seizure] = '2' or [seizure] = '3'

When did the seizures/epilepsy begin?	radio

1	Before age 30 years

2	Age 30 years or older

3	Don't know

53	other_preexist_yn	Do they have other pre-existing conditions not listed above?	radio

1	Yes

2	No

3	Don't know

54	other_preexist_des

Show the ﬁeld ONLY if: [other_preexist_yn] = '1'

Please list other conditions that are currently diagnosed.	text

| 1 | Yes |
| --- | --- |
| 2 | No |
| 3 | Don't know |

| 55 | other_meds | Section Header:  Are they currently being treated with any of the following medications for their pre-existing conditions (check all that apply)? | checkbox | | | | |
| --- | --- | --- | --- | --- | --- | --- | --- |
|  |  |  |  | 1 | other_meds_ 1 | | Angiotensin-converting enzyme (ACE) inhibitors (examples are: Benazepril (Lotensin), Captopril, Enalapril (Vasotec), Fosinopril, Lisinopril (Prinivil, Zestril), Moexipril, Perindopril, Quinapril (Accupril), Ramipril (Altace), Trandolapril)) |
|  |  |  |  | 2 | other_meds_ 2 | | Angiotensin receptor blockers (examples are: Azilsartan (Edarbi), Candesartan (Atacand), Eprosartan, Irbesartan (Avapro), Losartan (Cozaar), Olmesartan (Benicar), Telmisartan (Micardis), Valsartan (Diovan)) |
|  |  |  |  | 3 | other_meds_ 3 | | Systemic steroids given by mouth or injection (examples are: hydrocortisone (Cortef), cortisone, ethamethasoneb (Celestone), prednisone (Prednisone Intensol), prednisolone (Orapred, Prelone)) |
|  |  |  |  | 4 | other_meds_ 4 | | Immunosuppressants (e.g. anti-cancer or anti-rejection treatment) |
|  |  |  |  | 7 | other_meds_ 7 | | Palivizumab (sometimes given to young children who have problems associated with prematurity, lung or heart disease) |
|  |  |  |  | 5 | other_meds_ 5 | | Other |
|  |  |  |  | 6 | other_meds_ 6 | | None of the above |
|  |  |  | Field Annotation: @NONEOFTHEABOVE=6 | | | | |
| 56 | more_meds  Show the ﬁeld ONLY if: [other_meds(5)] = '1' | Please specify other medications for pre-existing conditions  (separate each medication by a comma). | text | | | | |
| 57 | vac_yn_fe | Section Header:  Do you know if the person with Down syndrome was  vaccinated for preventable illnesses in the past or currently (for example, chicken pox or the seasonal ﬂu)? | radio | | | | |
|  |  |  |  | 1 | Yes |  | |
|  |  |  |  | 2 | No |  |  |
|  |  |  |  | 3 | Don't know |  |  |
| 58 | ﬂu_fe  Show the ﬁeld ONLY if: [vac_yn_fe] = '1' | Seasonal ﬂu (inﬂuenza) vaccine -- given within the last 12-18 months | radio (Matrix) | | | | |
|  |  |  |  | 1 | Yes |  | |
|  |  |  |  | 2 | No |  |  |
|  |  |  |  | 3 | Don't know |  |  |
| 59 | td_fe  Show the ﬁeld ONLY if: [vac_yn_fe] = '1' | Tetanus, diphtheria, and pertussis (Td or Tdap) | radio (Matrix) | | | | |
|  |  |  |  | 1 | Yes |  | |
|  |  |  |  | 2 | No |  |  |
|  |  |  |  | 3 | Don't know |  |  |
| 60 | cp_fe  Show the ﬁeld ONLY if: [vac_yn_fe] = '1' | Chicken pox (Varicella, VZV) | radio (Matrix) | | | | |

61	mmr_fe

Show the ﬁeld ONLY if: [vac_yn_fe] = '1'

Measles mumps rubella vaccine (MMR)	radio (Matrix)

1	Yes

2	No

3	Don't know

62	pol_fe

Show the ﬁeld ONLY if: [vac_yn_fe] = '1'

Polio vaccine	radio (Matrix)

1	Yes

2	No

3	Don't know

63	rsv_fe

Show the ﬁeld ONLY if: [vac_yn_fe] = '1'

Respiratory syncytial virus (RSV) prophylaxis prophylaxis	radio (Matrix)

1	Yes

2	No

3	Don't know

64	shinnew_fe

Show the ﬁeld ONLY if: [vac_yn_fe] = '1'

Shingles -- Shingrix (newer, 2-dose vaccine)	radio (Matrix)

1	Yes

2	No

3	Don't know

65	shinold_fe

Show the ﬁeld ONLY if: [vac_yn_fe] = '1'

Shingles -- Zostavax (older vaccine)	radio (Matrix)

1	Yes

2	No

3	Don't know

66	pne_fe

Show the ﬁeld ONLY if: [vac_yn_fe] = '1'

Pneumococcal vaccine	radio (Matrix)

1	Yes

2	No

3	Don't know

67	hib_fe

Show the ﬁeld ONLY if: [vac_yn_fe] = '1'

Haemophilus inﬂuenza vaccine (Hib)	radio (Matrix)

1	Yes

2	No

3	Don't know

68	tub_fe

Show the ﬁeld ONLY if: [vac_yn_fe] = '1'

Tuberculosis vaccine (Bacille Calmette-Guerin, BCG)	radio (Matrix)

1	Yes

2	No

3	Don't know

69	yf_fe

Show the ﬁeld ONLY if: [vac_yn_fe] = '1'

Yellow fever vaccine (YF)	radio (Matrix)

1	Yes

2	No

3	Don't know

70	rot_fe

Show the ﬁeld ONLY if: [vac_yn_fe] = '1'

Severe diarrhea vaccine (Rotavirus vaccine)	radio (Matrix)

1	Yes

2	No

3	Don't know

71	hepa_fe

Show the ﬁeld ONLY if: [vac_yn_fe] = '1'

| 1 | Yes |
| --- | --- |
| 2 | No |
| 3 | Don't know |

Hepatitis A vaccine (Havrix, Vaqta)	radio (Matrix)

1	Yes

2	No

3	Don't know

72	hepb_fe

Show the ﬁeld ONLY if: [vac_yn_fe] = '1'

Hepatitis B vaccine	radio (Matrix)

73	men_fe

Show the ﬁeld ONLY if: [vac_yn_fe] = '1'

Meningitis (Meningococcal) vaccine	radio (Matrix)

1	Yes

2	No

3	Don't know

74	hpv_fe

Show the ﬁeld ONLY if: [vac_yn_fe] = '1'

Papillomavirus (HPV) vaccine	radio (Matrix)

1	Yes

2	No

3	Don't know

75	oth_vac_fe

Show the ﬁeld ONLY if: [vac_yn_fe] = '1'

Other scheduled vaccinations	radio (Matrix)

1	Yes

2	No

3	Don't know

76	oth_vac_list_fe

Please list other vaccines	text

Show the ﬁeld ONLY if: [oth_vac_fe] = '1'

77	test_done_yn	Section Header: *COVID-19 testing*

Was COVID-19 testing performed (swab or blood test)?

radio

1	Yes

2	No

3	Don't know

78	testing_why

Show the ﬁeld ONLY if: [test_done_yn] = '1'

Why was COVID-10 testing done?	radio

1	Symptoms were present

2	Possible contact with others who were aﬀected but no symptoms as yet

3	Routine testing--no speciﬁc indication

4	Don't know

79	test_result

Show the ﬁeld ONLY if: [test_done_yn] = '1'

What were the results of the testing?	radio

1	Pending

2	Preliminary test positive, conﬁrmatory test pending

3	Preliminary test positive, conﬁrmatory test negative

4	Conﬁrmatory test positive

5	Test negative

6	Don't know

Field Annotation: @HIDDEN

80	test_swab

Show the ﬁeld ONLY if: [test_done_yn] = '1'

| 4 | Test not done |
| --- | --- |
| 1 | Test done: positive (antibodies detected) |
| 2 | Test done: negative (antibodies not detected) |
| 3 | Test done: results are pending |
| 5 | Don't know or results were ambiguous |

Was a nose / throat swab test performed?	radio

4	Test not done

1	Test done: positive result

2	Test done: negative result

3	Test done: results are pending

5	Don't know or results were ambiguous

81	sars_fe

Show the ﬁeld ONLY if: [test_done_yn] = '1'

Was a blood test performed to test for antibodies against

COVID-19?

radio

82	symp_yn	Section Header:

Do/did they have signs and symptoms of COVID-19?

radio

1	Yes

2	No

3	Don't know

83	symp_present

Show the ﬁeld ONLY if: [symp_yn] = '1'

Please check any of the following signs or symptoms related to

COVID-19 that they experienced.

checkbox

1	symp_present_ 1	Fever of 100.4°F/38°C or higher

2	symp_present_ 2	Chills

3	symp_present_ 3	Nasal signs (congestion, runny nose, etc)

15	symp_present_ 15	Cough

4	symp_present_ 4	Headache

5	symp_present_ 5	Sore throat

6	symp_present_ 6	Shortness of breath

7	symp_present_ 7	Nausea or vomiting

8	symp_present_ 8	Abdominal pain

9	symp_present_ 9	Diarrhea (3 or more loose/looser than normal stools within 24 hrs)

10	symp_present_ 10	Muscle or joint pain

17	symp_present_ 17	Skin lesions (example, rash, ulcers, chilblain-like lesions)

18	symp_present_ 18	Conjunctivitis (sometimes called "pink eye")

11	symp_present_ 11	Loss of smell or taste

16	symp_present_ 16	Not eating or drinking

12	symp_present_ 12	Extreme fatigue, confusion, diﬃculty staying alert

13	symp_present_ 13	Unexplained behavioral change

14	symp_present_ 14	Other

84	oth_symp

Show the ﬁeld ONLY if: [symp_present(14)] = '1'

Please specify.	text

85	skin_loc

Show the ﬁeld ONLY if: [symp_present(17)] = '1'

Location of skin lesions (Please check all that apply):	checkbox

1	skin_loc_ 1	Head and neck

2	skin_loc_ 2	Trunk

3	skin_loc_ 3	Upper limbs / hands

4	skin_loc_ 4	Lower limbs / toes

5	skin_loc_ 5	Mucous membranes

6	skin_loc_ 6	Other

86	skin_loc_spec

Please specify	text

Show the ﬁeld ONLY if: [symp_present(17)] = '1'

87	hospital_yn	Section Header:

Was the person with Down syndrome admitted to a hospital?

radio

1	Yes

2	No

3	Don't know

88	days

Show the ﬁeld ONLY if: [hospital_yn] = '1'

How many days in hospital?	text (number, Min: 1, Max: 1000)

| 89 | icu_yn  Show the ﬁeld ONLY if: [hospital_yn] = '1' | Were they in an intensive care unit (ICU)? | radio | | |
| --- | --- | --- | --- | --- | --- |
|  |  |  |  | 1 | Yes |
|  |  |  |  | 2 | No |
|  |  |  |  | 3 | Don't know |
| 90 | days_icu  Show the ﬁeld ONLY if: [icu_yn] = '1' | How many days in ICU? | text (number, Min: 1, Max: 1000) | | |
| 91 | ventilation  Show the ﬁeld ONLY if: [hospital_yn] = '1' | Were they put on a mechanical ventilation? | radio | | |
|  |  |  |  | 1 | Yes |
|  |  |  |  | 2 | No |
|  |  |  |  | 3 | Don't know |
| 92 | outcome | Section Header:  At the last evaluation, what was their clinical situation? | radio | | |
|  |  |  |  | 1 | Not currently in hospital but with symptoms |
|  |  |  |  | 2 | Currently in hospital with symptoms |
|  |  |  |  | 3 | Tested positive but still no symptoms |
|  |  |  |  | 4 | Recovered from COVID-19 |
|  |  |  |  | 5 | Died |
|  |  |  |  | 6 | Other |
|  |  |  |  | 7 | Don't know |
| 93 | outcome_other  Show the ﬁeld ONLY if: [outcome] = '6' | Please specify their clinical situation. | text | | |
| 94 | days_symp  Show the ﬁeld ONLY if: [outcome] = '1' or [outcome]  = '2' or [outcome] = '4' or [out come] = '5' or [outcome] = '6' | About how many days has/did the person with Down syndrome had/have symptoms of COVID-19? (Enter 0 if you do not know.) | text (integer, Min: 0, Max: 100) | | |
