## Supplementary material for "An international survey on the impact of COVID-19 in individuals with Down syndrome": Clinician survey

Emory University

Library Information Technology Services

**Down syndrome and COVID-19**

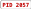

**Data Dictionary Codebook**

**Clinician Survey—English version**	10/25/2020 8:27am

% Collapse all instruments

| **#** | **Variable / Field Name** | **Field Label**  ***Field Note*** | **Field Attributes (Field Type, Validation, Choices, Calculations, etc.)** | | |
| --- | --- | --- | --- | --- | --- |
| Instrument: **Landing Page** (landing_page) ' Enabled as survey & Expand | | | | | |
| Instrument: **Family Eng** (family_eng) ' Enabled as survey & Expand | | | | | |
| Instrument: **Clinic Fu Eng** (clinic_fu_eng) ' Enabled as survey & Expand | | | | | |
| Instrument: **Clinic Sa Eng** (clinic_sa_eng) ' Enabled as survey % Collapse | | | | | |
| 217 | relation_cs | Section Header: *Please describe yourself and then answer general questions about the person with Down syndrome. Please write in "DK" or click on "Don't know" if you don't have the information.*  What is your relationship to the person with Down syndrome? | radio | | |
|  |  |  |  | 1 | Primary care physician |
|  |  |  |  | 2 | Clinician only caring for this person during  COVID-19 illness |
|  |  |  |  | 3 | Other |
| 218 | other_relation_cs  Show the ﬁeld ONLY if: [relation_cs] = '3' | Please specify your relationship. | text | | |
| 219 | age_cs | How old (in years) is the person with Down syndrome? (Please enter "0" if the person is a newborn baby.) | text (number, Min: 0, Max: 100), Required | | |
| 220 | gender_cs | How do they identify themselves? | radio, Required | | |
|  |  |  |  | 1 | Female |
|  |  |  |  | 2 | Male |
|  |  |  |  | 3 | Other |
|  |  |  |  | 4 | Choose not to answer |
| 221 | race_cs | What is their race? | text  Field Annotation: @HIDDEN | | |

| 1 | Borderline/normal |
| --- | --- |
| 2 | Mild |
| 3 | Moderate |
| 4 | Severe/Profound |
| 5 | Don't know |

| 222 | who_group_cs | What is their ethnic group (as deﬁned by World Health  Organization)? Please check all that apply: | checkbox | | | | | |
| --- | --- | --- | --- | --- | --- | --- | --- | --- |
|  |  |  |  | 1 | | who_group_cs 1 | | Aboriginal First Nations |
|  |  |  |  | 2 | | who_group_cs 2 | | Arab |
|  |  |  |  | 3 | | who_group_cs 3 | | Black |
|  |  |  |  | 4 | | who_group_cs 4 | | East Asian (e.g., China, Korea, Japan) |
|  |  |  |  | 5 | | who_group_cs 5 | | Latin American (e.g., Mexico, Brazil, Peru) |
|  |  |  |  | 6 | | who_group_cs 6 | | South Asian (e.g., Afghanistan, India, Pakistan) |
|  |  |  |  | 7 | | who_group_cs 7 | | West Asian (e.g., Saudi  Arabia, Syria, Turkey) |
|  |  |  |  | 8 | | who_group_cs 8 | | White |
|  |  |  |  | 9 | | who_group_cs 9 | | Other |
|  |  |  |  | 10 | | who_group_cs 10 | | Choose not to answer |
|  |  |  | Field Annotation: @NONEOFTHEABOVE=10 | | | | | |
| 223 | ethnicity_cs | How do they describe their ethnicity? | text | | | | | |
| 224 | trisomy_type_cs | Type of trisomy 21 | radio | | | | | |
|  |  |  |  | 1 | Full/standard | |  | |
|  |  |  |  | 2 | Mosaic | |  |  |
|  |  |  |  | 3 | Translocation | |  |  |
|  |  |  |  | 4 | Partial trisomy | |  |  |
|  |  |  |  | 5 | Don't know | |  |  |
| 225 | id_level_cs | Level of intellectual disability | radio  Field Annotation: @HIDECHOICE="1" | | | | | |

192	Yemen

193	Zambia

194	Zimbabwe

227	region_cs	What region of that country (e.g., state, province, district, county)?

text

228	setting_yn	Do you know the living situation of the person with Down syndrome before their illness or testing positive for COVID-19 (e.g., they live with their family, etc)?

radio

1	Yes

2	No

229	residence_cs

Show the ﬁeld ONLY if: [setting_yn] = '1'

What best describes their living situation before the COVID-19 outbreak?

radio

Field Annotation: @HIDECHOICE="6"

| 1 | Living at home with family |
| --- | --- |
| 7 | Living alone with no support (autonomously) |
| 2 | Living alone with support |
| 3 | Small group home with support |
| 4 | Residential care facility |
| 5 | Other |
| 6 | Don't know |

230	residence_other_cs

Show the ﬁeld ONLY if: [residence_cs] = '5'

Please specify other setting.	text

231	change_residence_cs

Show the ﬁeld ONLY if: [setting_yn] = '1'

Did they change their living situation because of the COVID-19 outbreak?

radio

1	Yes

2	No

3	Don't know

232	change_cs

Show the ﬁeld ONLY if: [change_residence_cs] = '1'

Please describe their current living situation.	radio

1	Living at home with family

7	Living alone with no support (autonomously)

2	Living alone with support

3	Small group home with support

4	Residential care facility

5	Other

6	Don't know / Not applicable

233	change_oth_cs

Show the ﬁeld ONLY if: [change_cs] = '5'

Please describe the other setting.	text

234	contact_yn_cs	Did anyone test positive or have signs or symptoms of

COVID-19 where the person with Down syndrome lived during the outbreak?

radio

1	Yes

2	No

3	Don't know / Not applicable

235	pre_exist_yn_cs	Section Header: *This section asks about medical conditions that existed before the COVID-19 outbreak (referred to as "pre-existing conditions").*

Can you provide information on pre-existing conditions?

| 1 | Yes |
| --- | --- |
| 2 | No |
| 3 | Don't know |

radio

1	Yes

2	No

236	chd_yn_cs

Show the ﬁeld ONLY if: [pre_exist_yn_cs] = '1'

Did the person with Down syndrome have a congenital heart defect?

radio

237	chd_surgery_cs

Show the ﬁeld ONLY if: [chd_yn_cs] = '1'

Did the heart defect require surgery?	radio

1	Yes surgery was done and fully correct the defect

2	Yes surgery was done but did not fully correct the defect

3	No surgery was required

4	Don't know

238	psych_yn_cs

Show the ﬁeld ONLY if: [pre_exist_yn_cs] = '1'

Does the person with Down syndrome have a current diagnosis of any behavioral and psychiatric condition (for example,

autism spectrum disorder)?

radio

1	Yes

2	No

3	Don't know

239	psych_type_cs

Show the ﬁeld ONLY if: [psych_yn_cs] = '1'

Please check all that apply.	checkbox

1	psych_type_cs_	1	Autism spectrum disorder

2	psych_type_cs_	2	Depression

3	psych_type_cs_	3	ADHD/ADD

4	psych_type_cs_	4	Anxiety

5	psych_type_cs_	5	Obsessive-compulsive disorder (OCD)

6	psych_type_cs_	6	Behavior problems

7	psych_type_cs_	7	Psychosis

8	psych_type_cs_	8	Other (regression disorder/disintegrative disorder, ...)

240	psych_other_cs

Please specify.	text

Show the ﬁeld ONLY if: [psych_type_cs(8)] = '1'

241	alz_cs

Show the ﬁeld ONLY if: [pre_exist_yn_cs] = '1'

Section Header: *Please check the following pre-existing conditions diagnosed by a doctor if experienced by the person with Down syndrome.*

Alzheimer disease/dementia

radio (Matrix)

1	Yes--currently being treated

2	Yes--not being treated

3	In the past--no current treatment

4	Never diagnosed

5	Don't know

242	thyroid_cs

Show the ﬁeld ONLY if: [pre_exist_yn_cs] = '1'

Thyroid disorder	radio (Matrix)

1	Yes--currently being treated

2	Yes--not being treated

3	In the past--no current treatment

4	Never diagnosed

5	Don't know

243	seizure_cs

Show the ﬁeld ONLY if: [pre_exist_yn_cs] = '1'

| 1 | Yes--currently being treated |
| --- | --- |
| 2 | Yes--not being treated |
| 3 | In the past--no current treatment |
| 4 | Never diagnosed |
| 5 | Don't know |

Seizures/epilepsy	radio (Matrix)

1	Yes--currently being treated

2	Yes--not being treated

3	In the past--no current treatment

4	Never diagnosed

5	Don't know

244	blood_cancer_cs

Show the ﬁeld ONLY if: [pre_exist_yn_cs] = '1'

Blood cancer (e.g., leukemia,lymphoma)	radio (Matrix)

245	other_cancer_cs

Show the ﬁeld ONLY if: [pre_exist_yn_cs] = '1'

Other cancer	radio (Matrix)

1	Yes--currently being treated

2	Yes--not being treated

3	In the past--no current treatment

4	Never diagnosed

5	Don't know

246	immumo_cs

Show the ﬁeld ONLY if: [pre_exist_yn_cs] = '1'

Immuno-compromised (e.g,, on cancer treatment)	radio (Matrix)

1	Yes--currently being treated

2	Yes--not being treated

3	In the past--no current treatment

4	Never diagnosed

5	Don't know

247	osa_cs

Show the ﬁeld ONLY if: [pre_exist_yn_cs] = '1'

Obstructive sleep apnea	radio (Matrix)

1	Yes--currently being treated

2	Yes--not being treated

3	In the past--no current treatment

4	Never diagnosed

5	Don't know

248	obesity_cs

Show the ﬁeld ONLY if: [pre_exist_yn_cs] = '1'

Obesity	radio (Matrix)

1	Yes--currently being treated

2	Yes--not being treated

3	In the past--no current treatment

4	Never diagnosed

5	Don't know

249	hyperten_cs

Show the ﬁeld ONLY if: [pre_exist_yn_cs] = '1'

Hypertension	radio (Matrix)

1	Yes--currently being treated

2	Yes--not being treated

3	In the past--no current treatment

4	Never diagnosed

5	Don't know

250	diabetes_cs

Show the ﬁeld ONLY if: [pre_exist_yn_cs] = '1'

Diabetes	radio (Matrix)

1	Yes--currently being treated

2	Yes--not being treated

3	In the past--no current treatment

4	Never diagnosed

5	Don't know

251	cvd_cs

Show the ﬁeld ONLY if: [pre_exist_yn_cs] = '1'

| 1 | Yes--currently being treated |
| --- | --- |
| 2 | Yes--not being treated |
| 3 | In the past--no current treatment |
| 4 | Never diagnosed |
| 5 | Don't know |

Cerebrovascular disease	radio (Matrix)

1	Yes--currently being treated

2	Yes--not being treated

3	In the past--no current treatment

4	Never diagnosed

5	Don't know

252	cad_cs

Show the ﬁeld ONLY if: [pre_exist_yn_cs] = '1'

Coronary heart disease	radio (Matrix)

253	renal_cs

Show the ﬁeld ONLY if: [pre_exist_yn_cs] = '1'

Chronic renal disease	radio (Matrix)

1	Yes--currently being treated

2	Yes--not being treated

3	In the past--no current treatment

4	Never diagnosed

5	Don't know

254	liver_cs

Show the ﬁeld ONLY if: [pre_exist_yn_cs] = '1'

Chronic liver disease	radio (Matrix)

1	Yes--currently being treated

2	Yes--not being treated

3	In the past--no current treatment

4	Never diagnosed

5	Don't know

255	lung_cs

Show the ﬁeld ONLY if: [pre_exist_yn_cs] = '1'

Chronic lung disease (e.g., asthma, emphysema, COPD)	radio (Matrix)

1	Yes--currently being treated

2	Yes--not being treated

3	In the past--no current treatment

4	Never diagnosed

5	Don't know

256	celiac_cs

Show the ﬁeld ONLY if: [pre_exist_yn_cs] = '1'

Celiac disease	radio (Matrix)

1	Yes--currently being treated

2	Yes--not being treated

3	In the past--no current treatment

4	Never diagnosed

5	Don't know

257	gerd_cs

Show the ﬁeld ONLY if: [pre_exist_yn_cs] = '1'

Gastroesophageal reﬂux (GERD)	radio (Matrix)

1	Yes--currently being treated

2	Yes--not being treated

3	In the past--no current treatment

4	Never diagnosed

5	Don't know

258	ibs_cs

Show the ﬁeld ONLY if: [pre_exist_yn_cs] = '1'

Irritable bowel syndrome (IBS)	radio (Matrix)

1	Yes--currently being treated

2	Yes--not being treated

3	In the past--no current treatment

4	Never diagnosed

5	Don't know

259	hep_b_cs

Show the ﬁeld ONLY if: [pre_exist_yn_cs] = '1'

| 1 | Before age 30 years |
| --- | --- |
| 2 | Age 30 years or older |
| 3 | Don't know |

Heaptitis B infection	radio (Matrix)

1	Yes--currently being treated

2	Yes--not being treated

3	In the past--no current treatment

4	Never diagnosed

5	Don't know

260	onset_seiz_cs

Show the ﬁeld ONLY if: [seizure_cs] = '1' or [seizure_c s] = '2' or [seizure_cs] = '3'

When did the seizures/epilepsy begin?	radio

| 1 | Yes, |
| --- | --- |
| 2 | No |
| 3 | Don't know |

| 261 | other_preexist_yn_cs  Show the ﬁeld ONLY if: [pre_exist_yn_cs] = '1' | Do they have other pre-existing conditions not listed above? | radio | | | | |
| --- | --- | --- | --- | --- | --- | --- | --- |
|  |  |  |  | 1 | Yes |  | |
|  |  |  |  | 2 | No |  |  |
|  |  |  |  | 3 | Don't know |  |  |
| 262 | other_preexist_des_cs  Show the ﬁeld ONLY if: [other_preexist_yn_cs] = '1' | Please list other conditions that are currently diagnosed. | text | | | | |
| 263 | other_meds_cs | Section Header:  Are they currently being treated with any of the following medications for their pre-existing conditions (check all that apply)? | checkbox | | | | |
|  |  |  |  | 1 | other_meds_cs_ 1 | | Angiotensin-converting enzyme (ACE) inhibitors (examples are: Benazepril (Lotensin), Captopril, Enalapril (Vasotec), Fosinopril, Lisinopril (Prinivil, Zestril), Moexipril, Perindopril, Quinapril (Accupril), Ramipril (Altace), Trandolapril)) |
|  |  |  |  | 2 | other_meds_cs_ 2 | | Angiotensin receptor blockers (examples are: Azilsartan (Edarbi), Candesartan (Atacand), Eprosartan, Irbesartan (Avapro), Losartan (Cozaar), Olmesartan (Benicar), Telmisartan (Micardis), Valsartan (Diovan)) |
|  |  |  |  | 3 | other_meds_cs_ 3 | | Systemic steroids given by mouth or injection (examples are: hydrocortisone (Cortef), cortisone, ethamethasoneb (Celestone), prednisone (Prednisone Intensol), prednisolone (Orapred, Prelone)) |
|  |  |  |  | 4 | other_meds_cs_ 4 | | Immunosuppressants (e.g. anti-cancer or anti-rejection treatment) |
|  |  |  |  | 7 | other_meds_cs_ 7 | | Palivizumab (sometimes given to young children who have problems associated with prematurity, lung or heart disease) |
|  |  |  |  | 5 | other_meds_cs_ 5 | | Other |
|  |  |  |  | 6 | other_meds_cs_ 6 | | None of the above |
|  |  |  | Field Annotation: @NONEOFTHEABOVE=6 | | | | |
| 264 | more_meds_cs  Show the ﬁeld ONLY if: [other_meds_cs(5)] = '1' | Please specify other medications for pre-existing conditions  (separate each medication by a comma). | text | | | | |
| 265 | vac_yn_cs | Section Header:  Do you know if the person with Down syndrome was  vaccinated for preventable illnesses in the past or currently (for example, chicken pox or the seasonal ﬂu)? | radio | | | | |
|  |  |  |  | 1 | Yes |  | |
|  |  |  |  | 2 | No |  |  |
|  |  |  |  | 3 | Don't know |  |  |
| 266 | ﬂu_cs  Show the ﬁeld ONLY if: [vac_yn_cs] = '1' | Seasonal ﬂu (inﬂuenza) vaccine -- given within the last 12-18 months | radio (Matrix) | | | | |

267	td_cs

Show the ﬁeld ONLY if: [vac_yn_cs] = '1'

Tetanus, diphtheria, and pertussis (Td or Tdap)	radio (Matrix)

1	Yes,

2	No

3	Don't know

268	cp_cs

Show the ﬁeld ONLY if: [vac_yn_cs] = '1'

Chicken pox (Varicella, VZV)	radio (Matrix)

1	Yes,

2	No

3	Don't know

269	mmr_cs

Show the ﬁeld ONLY if: [vac_yn_cs] = '1'

Measles mumps rubella vaccine (MMR)	radio (Matrix)

1	Yes,

2	No

3	Don't know

270	pol_cs

Show the ﬁeld ONLY if: [vac_yn_cs] = '1'

Polio vaccine	radio (Matrix)

1	Yes,

2	No

3	Don't know

271	rsv_cs

Show the ﬁeld ONLY if: [vac_yn_cs] = '1'

Respiratory syncytial virus (RSV) prophylaxis	radio (Matrix)

1	Yes,

2	No

3	Don't know

272	shinnew_cs

Show the ﬁeld ONLY if: [vac_yn_cs] = '1'

Shingles -- Shingrix (newer, 2-dose vaccine)	radio (Matrix)

1	Yes,

2	No

3	Don't know

273	shinold_cs

Show the ﬁeld ONLY if: [vac_yn_cs] = '1'

Shingles -- Zostavax (older vaccine)	radio (Matrix)

1	Yes,

2	No

3	Don't know

274	pne_cs

Show the ﬁeld ONLY if: [vac_yn_cs] = '1'

Pneumococcal vaccine	radio (Matrix)

1	Yes,

2	No

3	Don't know

275	hib_cs

Show the ﬁeld ONLY if: [vac_yn_cs] = '1'

Haemophilus inﬂuenza vaccine (Hib)	radio (Matrix)

1	Yes,

2	No

3	Don't know

276	tub_cs

Show the ﬁeld ONLY if: [vac_yn_cs] = '1'

Tuberculosis vaccine (Bacille Calmette-Guerin, BCG)	radio (Matrix)

1	Yes,

2	No

3	Don't know

277	yf_cs

Show the ﬁeld ONLY if: [vac_yn_cs] = '1'

| 1 | Yes, |
| --- | --- |
| 2 | No |
| 3 | Don't know |

Yellow fever vaccine (YF)	radio (Matrix)

1	Yes,

2	No

3	Don't know

278	rot_cs

Show the ﬁeld ONLY if: [vac_yn_cs] = '1'

Severe diarrhea vaccine (Rotavirus vaccine)	radio (Matrix)

279	hepa_cs

Show the ﬁeld ONLY if: [vac_yn_cs] = '1'

Hepatitis A vaccine (Havrix, Vaqta)	radio (Matrix)

1	Yes,

2	No

3	Don't know

280	hepb_cs

Show the ﬁeld ONLY if: [vac_yn_cs] = '1'

Hepatitis B vaccine	radio (Matrix)

1	Yes,

2	No

3	Don't know

281	men_cs

Show the ﬁeld ONLY if: [vac_yn_cs] = '1'

Meningitis (Meningococcal) vaccine	radio (Matrix)

1	Yes,

2	No

3	Don't know

282	hpv_cs

Show the ﬁeld ONLY if: [vac_yn_cs] = '1'

Papillomavirus (HPV) vaccine	radio (Matrix)

1	Yes,

2	No

3	Don't know

283	oth_vac_cs

Show the ﬁeld ONLY if: [vac_yn_cs] = '1'

Other scheduled vaccinations	radio (Matrix)

1	Yes,

2	No

3	Don't know

284	oth_vac_list_cs

Please list other vaccines	text

Show the ﬁeld ONLY if: [oth_vac_cs] = '1'

285	test_done_yn_cs	Section Header: *COVID-19 testing*

Was COVID-19 testing performed (swab or blood test)?

radio

1	Yes

2	No

3	Don't know

286	testing_why_cs

Show the ﬁeld ONLY if: [test_done_yn_cs] = '1'

Why was COVID-19 testing done?	radio

1	Symptoms were present

2	Possible contact with others who were aﬀected but no symptoms as yet

3	Routine testing--no speciﬁc indication

4	Don't know

287	test_result_cs

Show the ﬁeld ONLY if: [test_done_yn_cs] = '1'

What were the results of the testing?	radio

1	Pending

2	Preliminary test positive, conﬁrmatory test pending

3	Preliminary test positive, conﬁrmatory test negative

4	Conﬁrmatory test positive

5	Test negative

6	Don't know

Field Annotation: @HIDDEN

288	test_swab_cs

Show the ﬁeld ONLY if: [test_done_yn_cs] = '1'

Was a nasal / pharyngeal swab PCR test performed?	radio

| 4 | Test not done |
| --- | --- |
| 1 | Test done: positive result |
| 2 | Test done: negative result |
| 3 | Test done: results are pending |
| 5 | Don't know or results were ambiguous |

| 1 | skin_loc_cs_ 1 | Head and neck |
| --- | --- | --- |
| 2 | skin_loc_cs_ 2 | Trunk |
| 3 | skin_loc_cs_ 3 | Upper limbs / hands |
| 4 | skin_loc_cs_ 4 | Lower limbs / toes |
| 5 | skin_loc_cs_ 5 | Mucous membranes |
| 6 | skin_loc_cs_ 6 | Other |

| 289 | sars_cs  Show the ﬁeld ONLY if: [test_done_yn_cs] = '1' | Was a serology test performed to test for antibodies against  SARS-CoV-2? | radio | | | | | |
| --- | --- | --- | --- | --- | --- | --- | --- | --- |
|  |  |  |  | 4 | Test not done | | | |
|  |  |  |  | 1 | Test done: positive (antibodies detected) | | | |
|  |  |  |  | 2 | Test done: negative (antibodies not detected) | | | |
|  |  |  |  | 3 | Test done: results are pending | | | |
|  |  |  |  | 5 | Don't know or results were ambiguous | | | |
| 290 | symp_yn_cs | Section Header:  Do/did they have signs or symptoms of COVID-19? | radio | | | | | |
|  |  |  |  | 1 | Yes | |  | |
|  |  |  |  | 2 | No | |  |  |
|  |  |  |  | 3 | Don't know | |  |  |
| 291 | symp_present_cs  Show the ﬁeld ONLY if: [symp_yn_cs] = '1' | Please check any of the following symptoms related to  COVID-19 that they experienced. | checkbox | | | | | |
|  |  |  |  | 1 | | symp_present_cs_ 1 | | Fever of 100.4°F/38°C  or higher |
|  |  |  |  | 2 | | symp_present_cs_ 2 | | Chills |
|  |  |  |  | 3 | | symp_present_cs_ 3 | | Nasal signs (congestion, runny nose, etc) |
|  |  |  |  | 15 | | symp_present_cs_ 15 | | Cough |
|  |  |  |  | 4 | | symp_present_cs_ 4 | | Headache |
|  |  |  |  | 5 | | symp_present_cs_ 5 | | Sore throat |
|  |  |  |  | 6 | | symp_present_cs_ 6 | | Shortness of breath |
|  |  |  |  | 7 | | symp_present_cs_ 7 | | Nausea or vomiting |
|  |  |  |  | 8 | | symp_present_cs_ 8 | | Abdominal pain |
|  |  |  |  | 9 | | symp_present_cs_ 9 | | Diarrhea (3 or more loose/looser than normal stools within  24 hrs) |
|  |  |  |  | 10 | | symp_present_cs_ 10 | | Muscle or joint pain |
|  |  |  |  | 17 | | symp_present_cs_ 17 | | Skin lesions (example, rash, ulcers, chilblain- like lesions) |
|  |  |  |  | 18 | | symp_present_cs_ 18 | | Conjunctivitis (sometimes called "pink eye") |
|  |  |  |  | 11 | | symp_present_cs_ 11 | | Loss of smell or taste |
|  |  |  |  | 16 | | symp_present_cs_ 16 | | Not eating or drinking |
|  |  |  |  | 12 | | symp_present_cs_ 12 | | Extreme fatigue, confusion, diﬃculty staying alert |
|  |  |  |  | 13 | | symp_present_cs_ 13 | | Unexplained behavioral change |
|  |  |  |  | 14 | | symp_present_cs_ 14 | | Other |
| 292 | other_symp_cs  Show the ﬁeld ONLY if: [symp_present_cs(14)] = '1' | Please specify. | text | | | | | |
| 293 | skin_loc_cs  Show the ﬁeld ONLY if: [symp_present_cs(17)] = '1' | Location of skin lesions (Please check all that apply): | checkbox | | | | | |

294	skin_loc_spec_cs

Show the ﬁeld ONLY if: [skin_loc_cs(6)] = '1'

Please specify	text

295	skin_type_cs

Show the ﬁeld ONLY if: [symp_present_cs(17)] = '1'

Type of skin lesion (Please check all that apply):	checkbox

1	skin_type_cs_	1	Erythematous

2	skin_type_cs_	2	Maculopapules

3	skin_type_cs_	3	Vesicles

4	skin_type_cs_	4	Ulcer(s)

5	skin_type_cs_	5	Chilblain-like

6	skin_type_cs_	6	Other

296	skin_type_spec_cs

Please specify	text

Show the ﬁeld ONLY if: [skin_type_cs(6)] = '1'

297	hosp_yn_cs	Section Header:

Was the person with Down syndrome admitted to a hospital?

radio

1	Yes

2	No

3	Don't know

298	days_cs

Show the ﬁeld ONLY if: [hosp_yn_cs] = '1'

How many days in hospital?	text (number, Min: 1, Max: 1000)

299	icu_yn_cs

Show the ﬁeld ONLY if: [hosp_yn_cs] = '1'

Were they in an intensive care unit (ICU)?	radio

1	Yes

2	No

3	Don't know

300	days_icu_cs

How many days in ICU?	text (number, Min: 1, Max: 1000)

Show the ﬁeld ONLY if: [icu_yn_cs] = '1'

301	comp_yn_cs	Section Header:

Did they have medical complications due to COVID-19?

radio

1	Yes

2	No

3	Don't know

302	pneu_cs

Show the ﬁeld ONLY if: [comp_yn_cs] = '1'

Viral pneumonia associated with COVID-19	radio (Matrix)

1	Yes

2	No

3	Don't know

303	pneu_b_cs

Show the ﬁeld ONLY if: [comp_yn_cs] = '1'

Secondary bacterial pneumonia	radio (Matrix)

1	Yes

2	No

3	Don't know

304	respir_cs

Show the ﬁeld ONLY if: [comp_yn_cs] = '1'

| 1 | Yes |
| --- | --- |
| 2 | No |
| 3 | Don't know |

Acute respiratory distress syndrome	radio (Matrix)

1	Yes

2	No

3	Don't know

305	shock_cs

Show the ﬁeld ONLY if: [comp_yn_cs] = '1'

Septic shock	radio (Matrix)

306	kidney_cs

Show the ﬁeld ONLY if: [comp_yn_cs] = '1'

Acute kidney injury	radio (Matrix)

1	Yes

2	No

3	Don't know

307	coag_cs

Show the ﬁeld ONLY if: [comp_yn_cs] = '1'

Disseminated intravascular coagulation	radio (Matrix)

1	Yes

2	No

3	Don't know

308	throm_cs

Show the ﬁeld ONLY if: [comp_yn_cs] = '1'

Thrombotic complications	radio (Matrix)

1	Yes

2	No

3	Don't know

309	encep_cs

Show the ﬁeld ONLY if: [comp_yn_cs] = '1'

Encephalitis	radio (Matrix)

1	Yes

2	No

3	Don't know

310	rhab_cs

Show the ﬁeld ONLY if: [comp_yn_cs] = '1'

Rhabdomyolysis	radio (Matrix)

1	Yes

2	No

3	Don't know

311	cardiac_cs

Show the ﬁeld ONLY if: [comp_yn_cs] = '1'

Cardiac failure	radio (Matrix)

1	Yes

2	No

3	Don't know

312	hem_cs

Show the ﬁeld ONLY if: [comp_yn_cs] = '1'

Hemorrhage	radio (Matrix)

1	Yes

2	No

3	Don't know

313	organ_cs

Show the ﬁeld ONLY if: [comp_yn_cs] = '1'

Multiple organ dysfunction syndrome	radio (Matrix)

1	Yes

2	No

3	Don't know

314	misc_cs

Show the ﬁeld ONLY if: [comp_yn_cs] = '1'

Multisystem inﬂammatory syndrome in children (MIS-C) or

Kawasaki-like disease

radio (Matrix)

1	Yes

2	No

3	Don't know

315	other_comp_cs

Show the ﬁeld ONLY if: [comp_yn_cs] = '1'

Other	radio (Matrix)

1	Yes

2	No

3	Don't know

316	other_comp_sp_cs

Please specify.	text

Show the ﬁeld ONLY if: [other_comp_cs] = '1'

317	meds_yn_cs	Section Header:

Are/was the person treated with medications for COVID-19?

radio

| 1 | Yes |
| --- | --- |
| 2 | No |
| 3 | Don't know |

318	azyh_cs

Show the ﬁeld ONLY if: [meds_yn_cs] = '1'

Azithromycin	radio (Matrix)

1	Yes

2	No

3	Don't know

319	anti_biotic_cs

Show the ﬁeld ONLY if: [meds_yn_cs] = '1'

Other antibiotics (oral or IV)	radio (Matrix)

1	Yes

2	No

3	Don't know

320	chloro_cs

Show the ﬁeld ONLY if: [meds_yn_cs] = '1'

Chloroquine	radio (Matrix)

1	Yes

2	No

3	Don't know

321	hydro_cs

Show the ﬁeld ONLY if: [meds_yn_cs] = '1'

Hydroxychloroquine	radio (Matrix)

1	Yes

2	No

3	Don't know

322	remdes_cs

Show the ﬁeld ONLY if: [meds_yn_cs] = '1'

Remdesivir	radio (Matrix)

1	Yes

2	No

3	Don't know

323	antivir_cs

Show the ﬁeld ONLY if: [meds_yn_cs] = '1'

Other antiviral agents (lopinavir/ritonavir, darunavir/ritonavir

...)

radio (Matrix)

1	Yes

2	No

3	Don't know

324	gluco_cs

Show the ﬁeld ONLY if: [meds_yn_cs] = '1'

Systemic glucocorticoids	radio (Matrix)

1	Yes

2	No

3	Don't know

325	immune_cs

Show the ﬁeld ONLY if: [meds_yn_cs] = '1'

IV immune globulin	radio (Matrix)

1	Yes

2	No

3	Don't know

326	tocil_cs

Show the ﬁeld ONLY if: [meds_yn_cs] = '1'

Tocilizumab	radio (Matrix)

1	Yes

2	No

3	Don't know

327	fung_cs

Show the ﬁeld ONLY if: [meds_yn_cs] = '1'

Antifungal medication	radio (Matrix)

1	Yes

2	No

3	Don't know

328	hep_pro_cs

Show the ﬁeld ONLY if: [meds_yn_cs] = '1'

| 1 | Yes |
| --- | --- |
| 2 | No |
| 3 | Don't know |

Low molecular weight heparins (prophilactic dose)	radio (Matrix)

1	Yes

2	No

3	Don't know

329	hep_ther_cs

Show the ﬁeld ONLY if: [meds_yn_cs] = '1'

Low molecular weight heparins (therapeutic dose)	radio (Matrix)

330	anti_coag_cs

Show the ﬁeld ONLY if: [meds_yn_cs] = '1'

Other anti-coagulants (oral or IV)	radio (Matrix)

1	Yes

2	No

3	Don't know

331	colch_cs

Show the ﬁeld ONLY if: [meds_yn_cs] = '1'

Colchicine	radio (Matrix)

1	Yes

2	No

3	Don't know

332	melat_cs

Show the ﬁeld ONLY if: [meds_yn_cs] = '1'

Melatonin	radio (Matrix)

1	Yes

2	No

3	Don't know

333	oth_treat_meds_cs

Show the ﬁeld ONLY if: [meds_yn_cs] = '1'

Other	radio (Matrix)

1	Yes

2	No

3	Don't know

334	oth_treat_meds_sp_cs

Please list other relevant medications.	text

Show the ﬁeld ONLY if: [oth_treat_meds_cs] = '1'

335	oxy_cs	Section Header: *Other treatments used/in use during COVID-19 illness*

Oxygen therapy

radio (Matrix)

1	Yes

2	No

3	Don't know

336	cpap_cs	CPAP / BIPAP	radio (Matrix)

1	Yes

2	No

3	Don't know

337	vent_cs	Mechanical ventilation	radio (Matrix)

1	Yes

2	No

3	Don't know

338	memb_oxy_cs	Extracorporeal membrane oxygenation	radio (Matrix)

1	Yes

2	No

3	Don't know

339	renal_replace_cs	Continuous renal-replacement therapy	radio (Matrix)

1	Yes

2	No

3	Don't know

340	other_treat_cs	Other	radio (Matrix)

1	Yes

2	No

3	Don't know

341	oth_treat_sp_cs

Show the ﬁeld ONLY if: [other_treat_cs] = '1'

Please list other relevant treatments.	text

| 342 | outcome_cs | Section Header:  At the last evaluation, what was their clinical situation? | radio | | |
| --- | --- | --- | --- | --- | --- |
|  |  |  |  | 1 | Not currently in hospital but with symptoms |
|  |  |  |  | 2 | Currently in hospital with symptoms |
|  |  |  |  | 3 | Tested positive but still no symptoms |
|  |  |  |  | 4 | Recovered from COVID-19 |
|  |  |  |  | 5 | Died |
|  |  |  |  | 6 | Other |
|  |  |  |  | 7 | Don't know |
| 343 | outcome_other_cs  Show the ﬁeld ONLY if: [outcome_cs] = '6' | Please specify their clinical situation. | text | | |
| 344 | days_symp_cs  Show the ﬁeld ONLY if: [outcome_cs] = '1' or [outcom e_cs] = '2' or [outcome_cs] = '4  ' or [outcome_cs] = '5' or [outc ome_cs] = '6' | About how many days has/did the person with Down syndrome had/have symptoms of COVID-19? Enter 0 if you do not know. | text (integer, Min: 0, Max: 100) | | |
